# A Common Longitudinal Intensive Care Unit data Format (CLIF) to enable multi-institutional federated critical illness research

**DOI:** 10.1101/2024.09.04.24313058

**Authors:** Juan C. Rojas, Patrick G. Lyons, Kaveri Chhikara, Vaishvik Chaudhari, Sivasubramanium V. Bhavani, Muna Nour, Kevin G. Buell, Kevin D. Smith, Catherine A. Gao, Saki Amagai, Chengsheng Mao, Yuan Luo, Anna K Barker, Mark Nuppnau, Haley Beck, Rachel Baccile, Michael Hermsen, Zewei Liao, Brenna Park-Egan, Kyle A Carey, XuanHan, Chad H Hochberg, Nicholas E Ingraham, William F Parker

## Abstract

**Background:** Critical illness, or acute organ failure requiring life support, threatens over five million American lives annually. Electronic health record (EHR) data are a source of granular information that could generate crucial insights into the nature and optimal treatment of critical illness. However, data management, security, and standardization are barriers to large-scale critical illness EHR studies.

**Methods:** A consortium of critical care physicians and data scientists from eight US healthcare systems developed the Common Longitudinal Intensive Care Unit (ICU) data Format (CLIF), an open-source database format that harmonizes a minimum set of ICU Data Elements for use in critical illness research. We created a pipeline to process adult ICU EHR data at each site. After development and iteration, we conducted two proof-of-concept studies with a federated research architecture: 1) an external validation of an in-hospital mortality prediction model for critically ill patients and 2) an assessment of 72-hour temperature trajectories and their association with mechanical ventilation and in-hospital mortality using group-based trajectory models.

**Results:** We converted longitudinal data from 94,356 critically ill patients treated in 2020-2021 (mean age 60.6 years [standard deviation 17.2], 30% Black, 7% Hispanic, 45% female) across 8 health systems and 33 hospitals into the CLIF format, The in-hospital mortality prediction model performed well in the health system where it was derived (0.81 AUC, 0.06 Brier score). Performance across CLIF consortium sites varied (AUCs: 0.74-0.83, Brier scores: 0.06-0.01), and demonstrated some degradation in predictive capability. Temperature trajectories were similar across health systems. Hypothermic and hyperthermic-slow-resolver patients consistently had the highest mortality.

**Conclusions:** CLIF facilitates efficient, rigorous, and reproducible critical care research. Our federated case studies showcase CLIF’s potential for disease sub-phenotyping and clinical decision-support evaluation. Future applications include pragmatic EHR-based trials, target trial emulations, foundational multi-modal AI models of critical illness, and real-time critical care quality dashboards.

## INTRODUCTION

The intensive care unit (ICU) is an optimal setting for advanced clinical artificial intelligence (AI) projects because of voluminous longitudinal data and comprehensively captured outcome labels, demonstrated by exemplar deidentified electronic health record (EHR) databases such as MIMIC.^1–3^ AI applications in the ICU range from early warning systems for patient deterioration to programs designed to optimize resource allocation and personalize treatment recommendations.^4,5^ However, real-world ICU data science is often inefficient and difficult to scale because of challenges in acquiring, organizing, cleaning, and harmonizing EHR data. ICU EHR data are complex, highly temporally correlated, and subject to degradation through data capture and storage procedures designed for purposes other than research.^6^

Local EHR data repositories, or Electronic Data Warehouses (EDWs), are designed to maintain source data integrity and meet various institutional research and operational needs.^7^ EDWs often have unique idiosyncrasies, syntax, and data vocabularies which means extensive preprocessing is required before data can be analyzed for a specific use case.^8–11^ Established open-source common data models (CDMs), such as the Observational Medical Outcomes Partnership (OMOP),^12^ seek to address this data harmonization and standardization challenge for the entire EHR. However, the extract-transform-load (ETL) to a CDM is a major data engineering challenge, and data elements essential for the study of critical illness are not prioritized. Local CDM instances often completely omit granular critical illness data elements, such as ventilator settings for patients suffering from respiratory failure.^13,14^

To address these challenges and to support efficient and scalable data-driven critical care research, we developed the Common Longitudinal ICU Data Format (CLIF), a standardized data format for multi-center federated studies of critical illness. Recognizing that transforming raw data into analysis-ready structures is inherently domain-specific, we integrated the experiences and expertise of ICU clinician-scientists and data scientists, encoding this knowledge into a 1) clinically-driven entity-relationship model and a 2) minimum set of essential Common ICU Data Elements. Our overall objective was to create a robust critical illness research format that fully captures the dynamic clinical state of critical illness.

## METHODS

### CLIF Consortium Objectives and Process

We assembled a geographically diverse group of US-based physician-scientists and data scientists experienced in EHR-based clinical outcomes and AI research. Our guiding principles were: (1) efficient, clinically understandable data structures; (2) consistent and harmonizable data elements; (3) scalability and flexibility for future advancements; (4) federated analysis for collaborative research while maintaining data privacy and security; and (5) open-source development in line with the 2023 NIH Data Management and Sharing Policy and FAIR (Findable, Accessible, Interoperable, Reusable) data principles^15^.

We began weekly virtual meetings in July 2023 to develop operating procedures, terminologies, and quality control methods. We identified the practical challenges of using EHR data to study critical illness locally and across centers (**Table 1**) and addressed them when developing CLIF. CLIF is specifically designed for tasks like cohort discovery, temporal sequencing, and creating composite representations of clinical events, such as sepsis onset.

**Table 1.** Practical Challenges to EHR Data Science in the Hospital.

| Challenge | Description | Example |
| --- | --- | --- |
| **Complex longitudinal data with differing frequencies drawn from multiple sources** | Diverse domains such as vital signs, laboratory measurements, medications, and respiratory support require different data structures for representation and analysis. | Vital signs are frequently recorded during hospitalization (e.g. hourly) while laboratory results occur much less frequently and can be distributed across different record id numbers for the same patient (e.g., tests obtained in a clinic before the patient is referred to the emergency department or admitted). Microbiology tests are similarly recorded but are further complicated by the possibility of multiple observations per test (e.g., blood culture positive for multiple organisms) and nested antimicrobial susceptibility testing. |
| **Interdependent data** | Complex care processes are implicitly embedded in the presence, absence, frequency, and content of structured data. | Continuous neuromuscular blockade (recorded in the medication administration table) requires invasive mechanical ventilation (recorded in the respiratory flowsheet tables). |
| **Temporally-dependent data** |  | Sepsis onset is defined by complex temporal heuristics involving the sequencing and timing of antimicrobials, infectious tests, and abnormal physiology. |
| **Inefficient and inaccurate data capture** | Many bedside measurements (e.g., vital signs, respiratory parameters) require manual recording or human validation of automatically recorded data before they are available in the EHR. | Respiratory flowsheets often contain carryforward and copy/paste observations, leading to internal inconsistencies (e.g., patients recorded as receiving low-flow nasal oxygen and invasive ventilation simultaneously). |
| **Complex data storage** | ICU data storage is fragmented across different systems (e.g., ventilator data, laboratory systems, vital signs), making comprehensive data analysis challenging without sophisticated integration efforts. Diverse end-user needs and goals (e.g., operational quality reporting vs. clinical research) | ICU data on ventilator settings may be stored separately from laboratory results or vital signs, requiring complex data integration for analysis. Additionally, the "one size fits all" approach in data warehouses can result in data formats that are not ideal for |
|  | lead enterprise and research data warehouses to adopt a “one size fits all” content and format approach for EHR data, which may not be optimal for specific research or operational needs. | specific research tasks, such as temporal analyses or patient-specific interventions. |
| **Local idiosyncrasies** | ICU practices and data recording can vary significantly between institutions, with local protocols influencing how data is recorded and stored, leading to variability that complicates multicenter studies. | ICU triage decisions, such as when to escalate care to invasive ventilation, are often based on local protocols, which can differ significantly between hospitals, leading to challenges in generalizing study findings across different settings. |

### CLIF entity-relationship model

CLIF’s entity-relationship (ER) model aligns with how critical care researchers organize and analyze clinical data in practice (**Figure 1**). It organizes various clinical data into 23 clinically relevant longitudinal tables linked by patient and hospitalization. These tables are organized by clinical information type and organ systems.

**Figure 1.**
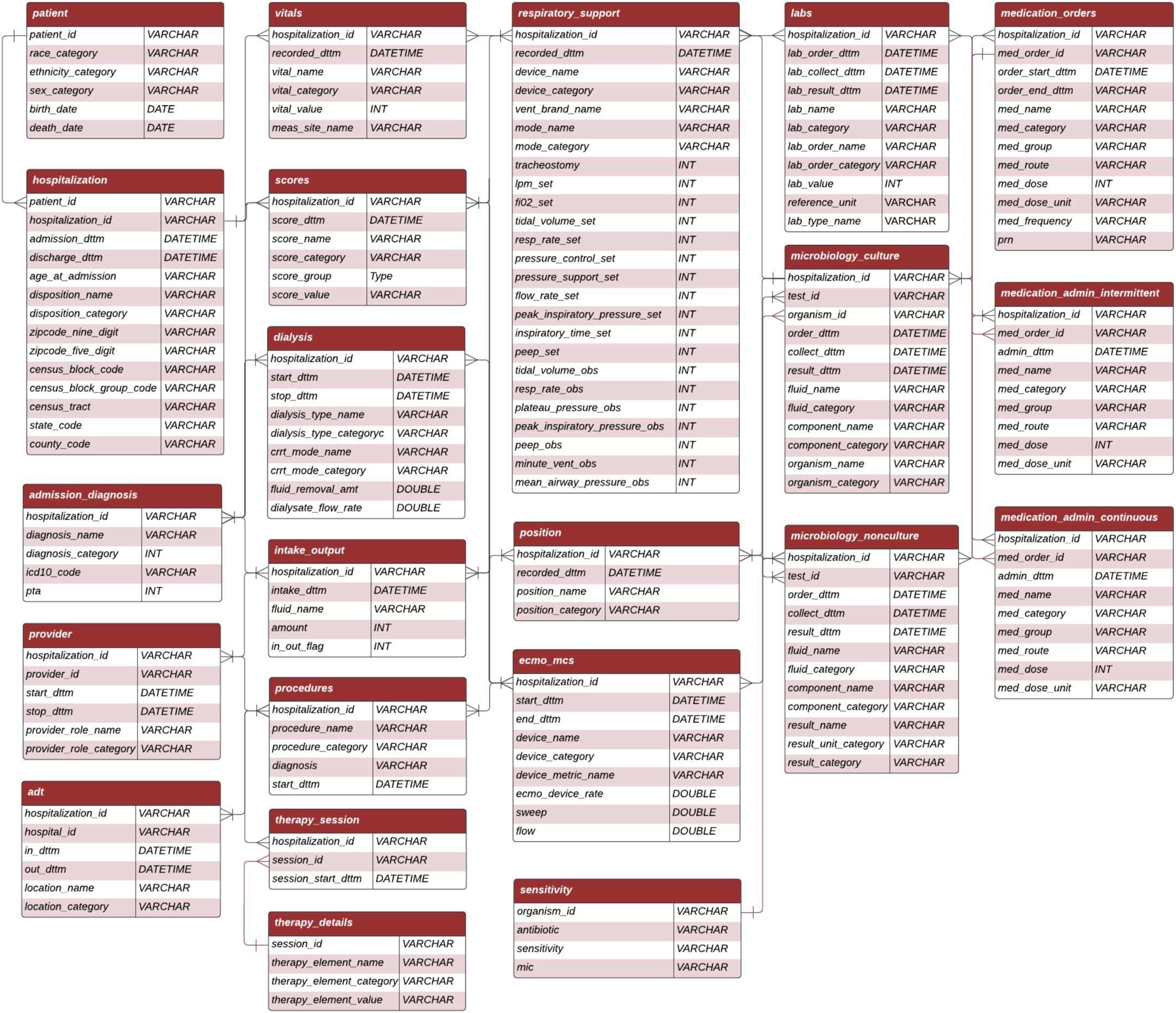
CLIF Entity Relationship Diagram. This diagram depicts the relationships between various tables in the Common Longitudinal ICU Data Format (CLIF) schema. The diagram includes the following 23 tables:

1. Patient
2. Hospitalization
3. Admission Diagnosis
4. Provider
5. ADT (Admission, Discharge, Transfer)
6. Vitals
7. Scores
8. Dialysis
9. Intake/Output
10. Procedures
11. Therapy session
12. Therapy Details
13. Respiratory Support
14. Position
15. ECMO (Extracorporeal Membrane Oxygenation) and Mechanical Circulatory Support (MCS)
16. Labs
17. Microbiology culture
18. Sensitivity
19. Microbiology non-culture
20. Respiratory Support
21. Medication Orders
22. Medication Admin Intermittent
23. Medication Admin Continuous Each table represents a specific aspect of ICU data, and lines between tables indicate how they are related through shared identifiers, primarily encounter_id. The depicted entity-relationship model was version 1.0, used for the case studies in this manuscript. The CLIF format is maintained with the git version control system, release 2.0.0 is available at https://clif-consortium.github.io/website/

CLIF’s ER model features specialized critical care tables such as respiratory support, continuous medications, dialysis, extra-corporeal membrane oxygenation/mechanical circulatory support, position (designed to identify prone mechanical ventilation), and scores (containing important clinical assessments such as the Glasgow Coma Scale or Richmond Agitation-Sedation Scale). The ER model also contains other standard inpatient EDW tables (e.g. vitals, labs) and can be implemented as Structured Query Language (SQL) views or in other database structures, as CLIF is language agnostic.

### Minimum Common ICU Data Elements and Preservation of source EHR Data

The National Institutes of Health (NIH) defines a Common Data Elements (CDE) as a "standardized, precisely defined question, paired with a set of allowable responses, used systematically across different sites, studies, or clinical trials to ensure consistent data collection.”^16^ For each CLIF table, we developed a minimum set of Common ICU Data Elements (mCIDE) denoted with the “_category” suffix. Each CIDE 1) represents a precisely defined clinical entity relevant to critical illness and 2) has a limited set of permissible values.

Whenever possible, we adopted NIH-endorsed CDEs into CLIF.^17^ We created several novel CIDEs for CLIF, such as modes of mechanical ventilation (mode_category). Recognizing that our mCIDE is insufficient for all research purposes, we preserve source EHR data elements in “_name” fields.” For example, lab_name (e.g., "UCM_LAB HEMOGLOBIN - AUTOMATED") preserves the specific lab test name as used at the site, while lab_category (e.g., "Hemoglobin") maps this test to a specific permissible value of the lab_category CDE. This structure creates standardized ICU elements while preserving original data labels for quality control.

### CLIF open-source commitment, AI disclosure, and IRB approval

CLIF continues to mature via our collaborative development process supported by git version control and is released under the Apache 2.0 license to ensure open access and broad usage rights. Our consortium website and code repository (clif-consortium.github.io/website/) contains data dictionaries, ETL pipeline examples, quality control scripts, and complete analysis code for each case study. Each of the eight CLIF consortium sites independently received IRB approval to conduct observational studies or to build and/or quality-check a research EDW (see Supplement **Table E1** for IRB details). No patient-level data was shared between sites at any point. We used AI-assisted technologies, including large language models (LLMs), to edit the manuscript and code analysis scripts. The authors carefully reviewed and verified all AI-generated content to ensure accuracy and originality. All quoted material is properly cited.

### Cohort discovery and federated analytics in the CLIF consortium

We conducted two proof-of-concept case studies: (1) development and external validation of a novel multivariable AI ICU mortality prediction model and (2) external validation of a previously developed subphenotyping algorithm for temperature trajectories in critical illness.^18^ We used the same cohort-discovery script and the CLIF admission-discharge-transfer (ADT) and patient tables to identify all adults (≥18 years) admitted to an ICU within 48 hours of hospitalization and staying at least 24 hours, from January 1, 2020, to December 31, 2021, at each site across the consortium (**Figure E1**). We chose these inclusion criteria to identify the general ICU population, excluding patients who die shortly after ICU admission or are admitted to the ICU for non-critical reasons (e.g., to facilitate a procedure). Using standardized outlier handling scripts and consortium-defined outlier ranges, we removed clear data entry errors (e.g., a heart rate of 1,000 beats per minute).

### Case Study I: Development and External Validation of an In-Hospital Mortality Model for ICU Patients

Accurate and reliable hospital mortality predictions for critically ill patients may help clinical teams prioritize therapeutic interventions, facilitate more informed shared decision-making around goals of care, and optimize resource allocation within healthcare systems. Existing prediction models are limited by suboptimal accuracy and significant performance variation across hospitals and differential performance among vulnerable populations may exacerbate baseline inequities in access to (and quality of) critical care. ^1920–23^ In this case study, we developed and externally validated an AI model to predict hospital mortality using clinical data from the first 24 hours in the ICU.

We trained a light gradient boosted machine binary classifier (LightGBM)^24^ to predict in-hospital death on separate cohort of ICU admissions in CLIF format from Rush University Medical Center using data from 2019, 2022, and 2023, performing hyperparameter tuning through a grid search with 5-fold cross validation. We selected LightGBM for its high discrimination and its ability to handle missing data without the need for imputation or exclusion of cases with high levels of missingness.^24^ We selected 30 candidate predictors *a priori* from hours 0-24 in the ICU (**Table E2)** based on literature review, our clinical experience, and expected low levels of missingness. We then saved the prediction model object in python and the general LGBM TXT format to the shared publicly available consortium repository.

We then evaluated this model on the 2020-2021 cohort described above at Rush and all other CLIF sites using a federated approach with a common model evaluation script, the model object, and each site’s local CLIF database. To comprehensively assess the model’s generalizability, we applied the TRIPOD-AI checklist across all test sites.^25^ Our evaluation focused on three key aspects: discrimination using the area under the receiver operating characteristic curve (AUC), calibration using Brier scores and calibration plots, and clinical utility through decision curve analysis.^25,26^

### Case Study II: Temperature Trajectory Subphenotyping

Growing recognition of heterogeneity within critical illness syndromes has led to the emergence of algorithmic clinical subphenotyping as a means of generating new hypotheses for investigation, improving clinical prognostication, and characterizing heterogeneous treatment effects.^27^ Despite the potential value of these advances in precision medicine, subphenotyping models are rarely externally validated.^28^

In our second case study, we externally validated a previously-developed unsupervised model to subphenotype infected patients in the hospital according to longitudinal temperature trajectories.^18^ This approach uses group-based trajectory modeling and patient temperature trends over 72 hours to assign patient encounters into one of four mutually-exclusive subphenotypes: normothermic (NT), hypothermic (HT), hyperthermic fast-resolver (HFR), and hyperthermic slow-resolver (HSR). In 1- and 2-hospital studies of patients with undifferentiated suspected infection and COVID-19 (regardless of ICU status), these subphenotypes have demonstrated distinct immune and inflammatory profiles and differential outcomes, including ICU utilization and mortality.^29–31^ However, the temperature trajectory model has not previously been evaluated within a broader critically ill population.

We developed analysis scripts that standardized body temperature measurements during the first 72 hours of ICU admission and classified each patient into the temperature trajectory subgroup with the lowest sum of the mean squared errors between the patient’s observed temperature and the subphenotype’s reference trajectory. Finally, we assessed differences in patient characteristics by subphenotype and the association of subphenotypes with in-hospital mortality and receipt of invasive mechanical ventilation using multivariable logistic regression adjusted for patient age, sex, race, and ethnicity.

## RESULTS

To date, we have established CLIF databases at eight US health systems, comprising 33 unique hospitals and 94,356 ICU admissions (**Figure E2, Table 2**). Health system-level populations were similar in terms of age (mean 60.6 years overall [standard deviation 17.2], range 56.4 [18.5] to 63.2 [17.5]) and sex (45% female overall, range 41-47% female). As expected, each site’s population differed substantially by race, ranging from 7% to 66% Black. Clinical outcomes differed moderately across systems. Overall, patients received invasive mechanical ventilation in 35,789 encounters (38% overall, range 28-50%), and 8,920 patients died in the hospital (9.5% overall, range 7.4-13%).

**Table 2.**
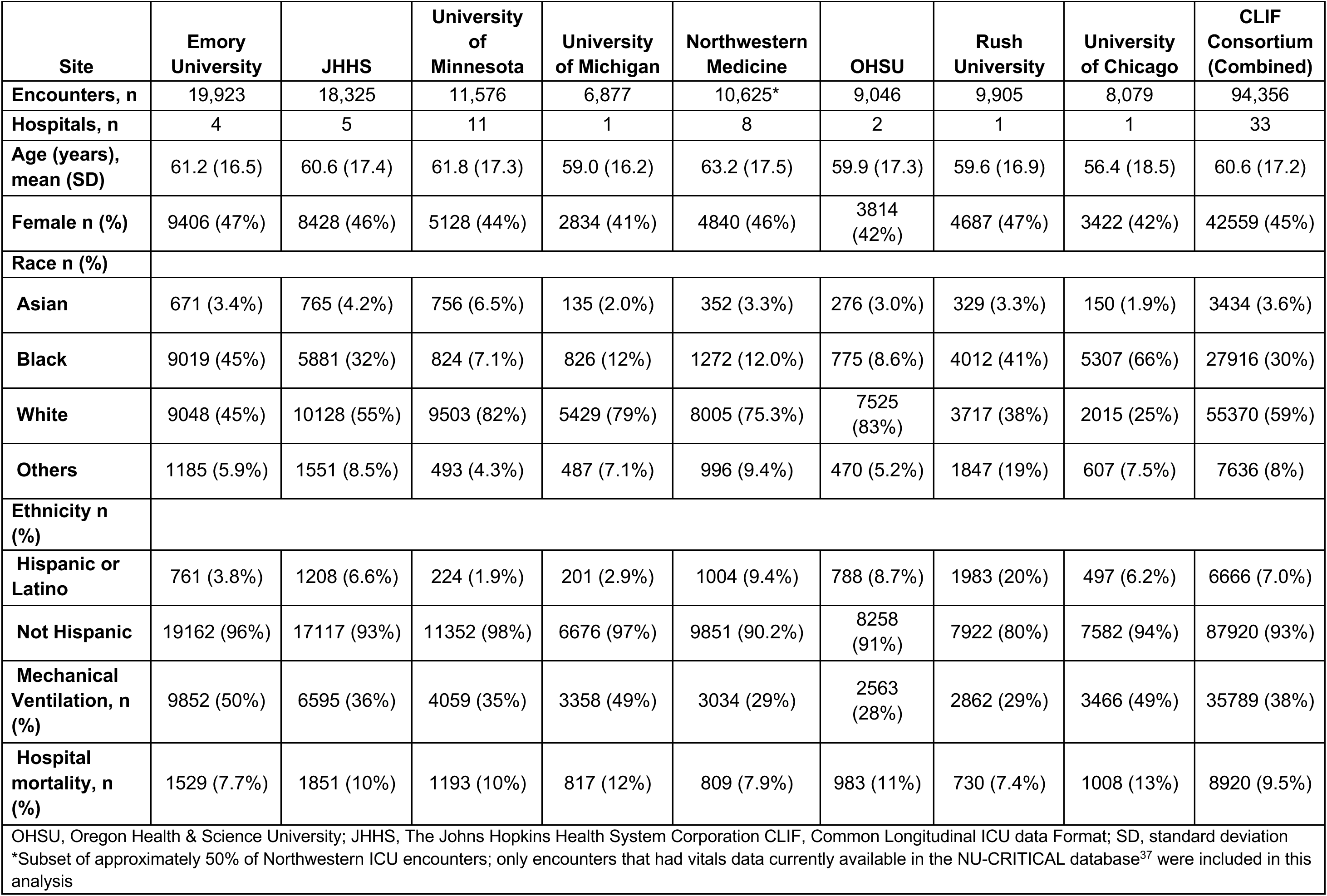
Characteristics and outcomes of ICU patient encounters in 2020-2021 across CLIF sites.

| Site | Emory University | JHHS | University of Minnesota | University of Michigan | Northwestern Medicine | OHSU | Rush University | University of Chicago | CLIF Consortium (Combined) |
| --- | --- | --- | --- | --- | --- | --- | --- | --- | --- |
| Encounters, n | 19,923 | 18,325 | 11,576 | 6,877 | 10,625* | 9,046 | 9,905 | 8,079 | 94,356 |
| Hospitals, n | 4 | 5 | 11 | 1 | 8 | 2 | 1 | 1 | 33 |
| Age (years), mean (SD) | 61.2 (16.5) | 60.6 (17.4) | 61.8 (17.3) | 59.0 (16.2) | 63.2 (17.5) | 59.9 (17.3) | 59.6 (16.9) | 56.4 (18.5) | 60.6 (17.2) |
| Female n (%) | 9406 (47%) | 8428 (46%) | 5128 (44%) | 2834 (41%) | 4840 (46%) | 3814 (42%) | 4687 (47%) | 3422 (42%) | 42559 (45%) |
| Race n (%) |  |  |  |  |  |  |  |  |  |
| Asian | 671 (3.4%) | 765 (4.2%) | 756 (6.5%) | 135 (2.0%) | 352 (3.3%) | 276 (3.0%) | 329 (3.3%) | 150 (1.9%) | 3434 (3.6%) |
| Black | 9019 (45%) | 5881 (32%) | 824 (7.1%) | 826 (12%) | 1272 (12.0%) | 775 (8.6%) | 4012 (41%) | 5307 (66%) | 27916 (30%) |
| White | 9048 (45%) | 10128 (55%) | 9503 (82%) | 5429 (79%) | 8005 (75.3%) | 7525 (83%) | 3717 (38%) | 2015 (25%) | 55370 (59%) |
| Others | 1185 (5.9%) | 1551 (8.5%) | 493 (4.3%) | 487 (7.1%) | 996 (9.4%) | 470 (5.2%) | 1847 (19%) | 607 (7.5%) | 7636 (8%) |
| Ethnicity n (%) |  |  |  |  |  |  |  |  |  |
| Hispanic or Latino | 761 (3.8%) | 1208 (6.6%) | 224 (1.9%) | 201 (2.9%) | 1004 (9.4%) | 788 (8.7%) | 1983 (20%) | 497 (6.2%) | 6666 (7.0%) |
| Not Hispanic | 19162 (96%) | 17117 (93%) | 11352 (98%) | 6676 (97%) | 9851 (90.2%) | 8258 (91%) | 7922 (80%) | 7582 (94%) | 87920 (93%) |
| Mechanical Ventilation, n (%) | 9852 (50%) | 6595 (36%) | 4059 (35%) | 3358 (49%) | 3034 (29%) | 2563 (28%) | 2862 (29%) | 3466 (49%) | 35789 (38%) |
| Hospital mortality, n (%) | 1529 (7.7%) | 1851 (10%) | 1193 (10%) | 817 (12%) | 809 (7.9%) | 983 (11%) | 730 (7.4%) | 1008 (13%) | 8920 (9.5%) |

OHSU, Oregon Health & Science University; JHHS, The Johns Hopkins Health System Corporation CLIF, Common Longitudinal ICU data Format; SD, standard deviation
\*Subset of approximately 50% of Northwestern ICU encounters; only encounters that had vitals data currently available in the NU-CRITICAL database<sup>37</sup> were included in this analysis

### Development and External Validation of an In-Hospital Mortality Model for ICU Patients

The separate Rush training cohort consisted of 17,139 ICU admissions with similar demographics to the Rush test cohort (**Table E3**). The final model hyperparameters of the lightGBM are described in the **Supplement (Table E4)**. The most important features in the model were: minimum albumin level, maximum aspartate aminotransferase (AST), minimum pulse rate, minimum diastolic blood pressure (DBP), and mean aspartate aminotransferase (AST) (see variable importance plot **Figure E3)**.

In the hold-out test cohort of 94,356 ICU admissions (**Table 1**), the AUC for predicting in-hospital mortality varied across sites, ranging from 0.74 to 0.81. Specifically, the Chicagoland health systems of Rush, Northwestern, and University of Chicago exhibited the highest AUCs, with values of 0.81 [95% CI: 0.79-0.83], 0.83 [0.81 - 0.84], and 0.81 [95% CI: 0.79-0.82], respectively. In contrast, Emory and the University of Minnesota reported the lowest AUCs, both at 0.74 [95% CI: 0.73-0.75] and 0.74 [95% CI: 0.72-0.75]. These performance metrics are summarized in **Figure 2a**. Calibration was assessed using Brier scores, which ranged from 0.059 at Oregon Health & Science University (OHSU) (indicating the best calibration) to 0.097 at the University of Michigan and the University of Chicago. The calibration plot (**Figure 2b**) demonstrates predicted versus observed probabilities across all sites, with most curves closely following the diagonal line at lower probabilities but some deviation at higher probabilities. Net benefit is a weighted average of true positives and false positives intended to quantify the clinical utility of a model at different treatment thresholds varied across sites, as shown in the decision-curve analysis in **Figure 2c**.^26^ At the high-risk threshold of a 0.3 probability of death determined *a priori* by Rush University, the model conferred the highest net benefit to the University of Chicago (0.025), followed by the University of Michigan (0.020), and John Hopkins (0.018). The model lead to the lowest net benefits at Emory and Oregon Health & Science University both at 0.006. The model had positive net benefit at all sites at the threshold, indicating it increased utility compared to a “treat none” strategy (net benefit = 0 by definition) and a “treat all” strategy (net benefit ranging from −0.24 to −0.32).

**Figure 2.**
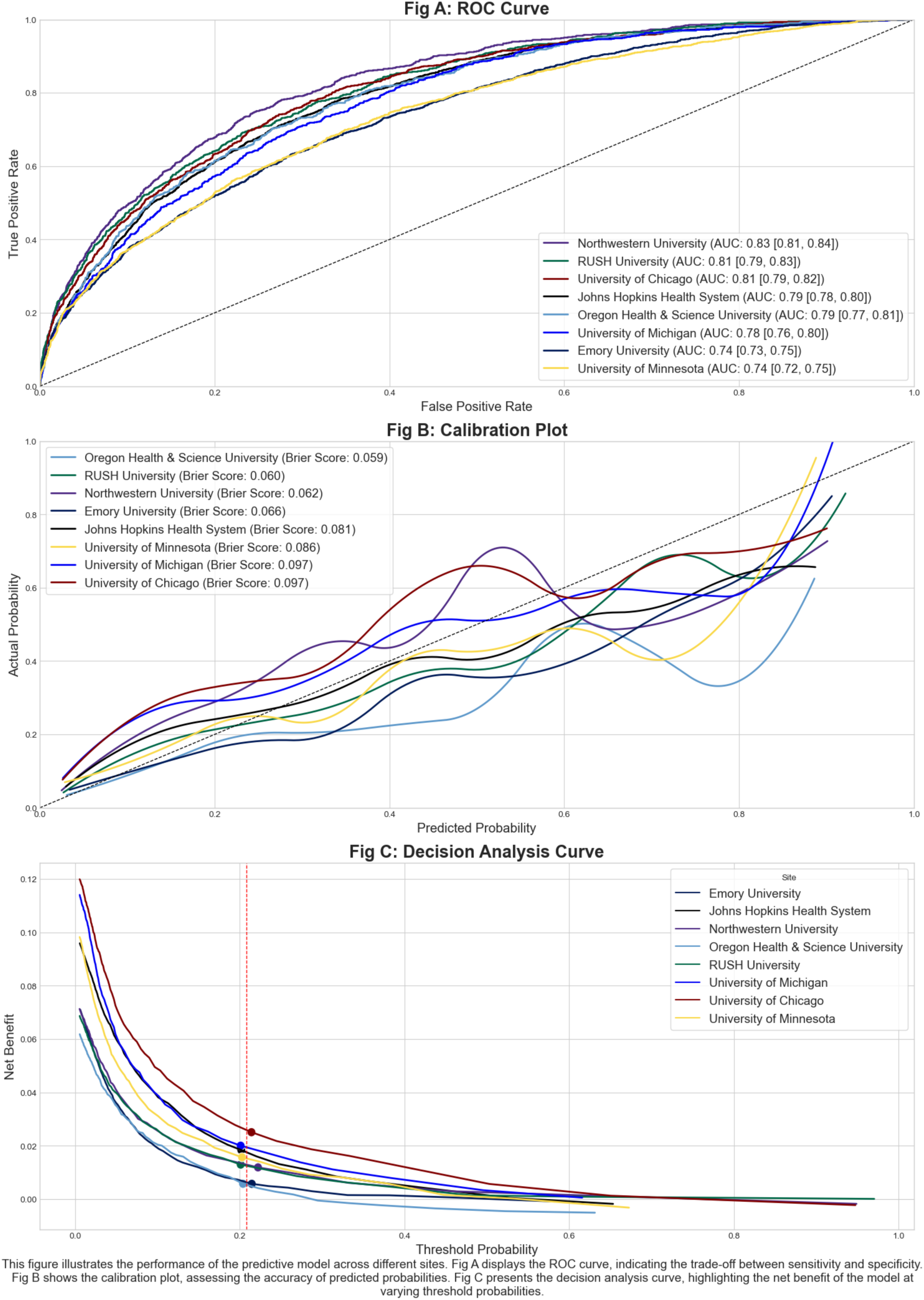
Mortality model performance validation via receiver-operator characteristic curves (A), calibration curves (B), and decision curves (C).

### Temperature Trajectory Subphenotypes

Across the eight participating institutions, this case study analyzed 94,290 ICU admissions, excluding 66 (0.07%) ICU admissions without any recorded temperatures (**Table E4**). Each subphenotype had a consistent observed temperature trajectory across all sites (**Figure 3A**). Normothermic encounters had the highest prevalence overall and at each site (range 43-68%), followed by hypothermic encounters (range 13-34%), hyperthermic slow resolvers (range 7.8-16%), and hyperthermic fast resolvers (range 6.2-7.9%). The distribution of patient characteristics across subphenotypes was similar across sites. Hypothermic patients were consistently older than other groups (range 61.4-68.9) and hyperthermic slow resolvers were the youngest group at all sites but one (range 47.8-60.0).

**Figure 3.**
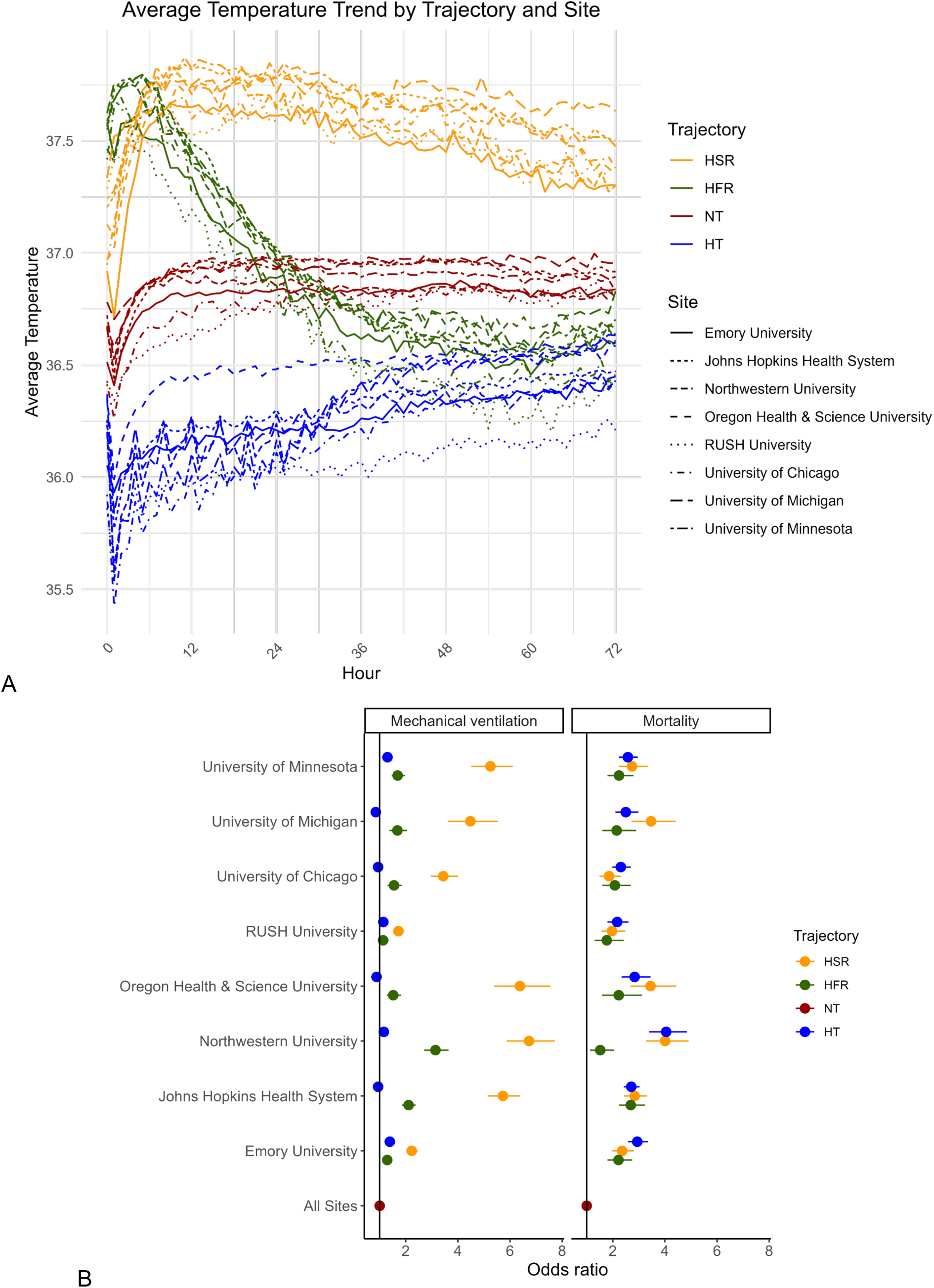
Temperature trends (A) and adjusted odds for ICU outcomes (B) across temperature trajectory subphenotypes at all sites.

Several consistent outcome patterns were observed across sites. Hyperthermic slow resolvers had the highest rates of invasive mechanical ventilation at all sites (range 38-79%). Mechanical ventilation rates were lowest among normothermic and hypothermic patients. Mortality was lowest among normothermic patients (range 3.8-8.2%) and highest among hypothermic (9.9-18.2%) and hyperthermic slow resolving (8.5-20.3%) patients.

After adjustment for age, sex, race, and ethnicity, subphenotype membership was consistently and independently associated with these outcomes (**Figure 3B**). HSR and HFR subphenotypes were associated with significantly increased odds for invasive mechanical ventilation (IMV) (as compared to normothermic) at all sites. Additionally, HSR, HFR, and HT subphenotypes were associated with significantly increased odds for mortality at all sites.

## DISCUSSION

We developed CLIF to standardize complex ICU data into a consistent, longitudinal format necessary for transparent and reproducible critical care research. In two proof-of-concept studies involving nearly 100,000 diverse critically ill patients, we demonstrated the potential of CLIF and a federated consortium research approach.

In our first case study, the mortality model demonstrated good discrimination and calibration in the internal RUSH validation cohort. However, its performance varied across seven other CLIF consortium sites. Decision curve analysis revealed positive but varying clinical utility across sites, highlighting the model’s sensitivity to local clinical and operational differences. For example, significant calibration slope errors (overestimating mortality) in the OHSU and Emory test sets led to lower net benefit and less clinical utility at the specified clinical decision threshold. These findings underscore the challenge of generalizing prognostic models across diverse healthcare settings in a one-size-fits-all fashion.^19^ This case study demonstrates CLIF’s value in facilitating rigorous, multi-site evaluations of predictive models. The natural next step is the development and validation of a set of generalizable ICU prediction models trained across the entire CLIF consortium using decentralized federated learning.^32^

Our second case study expands Bhavani et al.’s temperature trajectory subphenotyping model to a larger, broader, and more diverse cohort using the CLIF framework.^18^ While Bhavani et al. identified four subphenotypes in a sepsis-specific cohort—hyperthermic slow resolvers, hyperthermic fast resolvers, normothermic, and hypothermic—we applied this model to undifferentiated patients with critical illness.^18^ Mirroring prior findings in sepsis, we observed the highest mortality rates in the hypothermic group, suggesting this subphenotype robustly predicts outcomes in a general critical illness population. The consistent association of temperature trajectories with mortality and mechanical ventilation across health systems highlights the potential of longitudinal data analysis for critical illness phenotyping and personalizing ICU treatment.^33^

### Limitations and areas for improvement

The CLIF format has limitations and we are actively seeking feedback on the project. First, CLIF required substantial data science and critical care expertise to implement at each consortium site. The consortium is developing open-source tools to help diverse healthcare institutions adopt CLIF in an automated, scalable, and flexible manner. These ETL tools must be sensitive to the evolving landscape of healthcare data storage as institutions shift from traditional warehouses to flexible, scalable data lakes. This change allows CLIF to play a key role in improving healthcare data representation. CLIF helps transform often hard-to-interpret raw data in the ICU into more valuable, integrated information ready for advanced analysis. Second, CLIF is currently not linked to established interoperability standards like Health Level Seven (HL7) Fast Healthcare Interoperability Resources (FHIR). Future data engineering directions include the development of HL7 FHIR queries to generate real-time CLIF tables.

Third, the case studies presented in this manuscript used ADT data to define the critical illness population. This approach is susceptible to selection bias and left-censoring. However, a key strength of the CLIF framework is it is designed to represent critical illness wherever it occurs in the hospital.^34^ This means that future CLIF analyses do not have to rely on a patient’s physical location or ADT data to define a disease state. Fourth, while CLIF’s diverse data elements are not a substitute for proper longitudinal study design. Limitations like incomplete data or collider bias can still confound CLIF studies, especially when hospitalization patterns vary among vulnerable groups.^35^ Fortunately, advances in causal inference, such as target trial emulation, provide a clear roadmap for the CLIF consortium.

Finally, while our work demonstrates the benefits of federated analysis, ideally, we would release de-identified versions of our CLIF databases for public use. Once CLIF’s utility as a format is firmly established, we hope to make the case to our health systems leadership to make the large investment required to follow the inspirational example of MIMIC.^36^

### Conclusions and future directions

We developed and implemented a Common Longitudinal ICU data Format (CLIF) across 8 diverse health systems and demonstrated its value in two proof-of-concept case studies. We believe our open-source approach will make CLIF a broadly appealing target format for representing critical illness. Aspirational future directions for CLIF include 1) data format for pragmatic EHR-based trials, 2) rigorous target trial emulation framework for causal inference, 3) real-time critical care quality dashboards, and 4) foundational multi-modal AI models of critical illness.

## Data Availability

All aggregate results from the paper and prediction model objects are available online at https://clif-consortium.github.io/website/.

While patient row-level data cannot be shared due to patient privacy concerns, the authors will run are available via federated analysis using the CLIF format upon reasonable request.

https://clif-consortium.github.io/website/

## Supplementary Tables and Figures

**Table E1:**
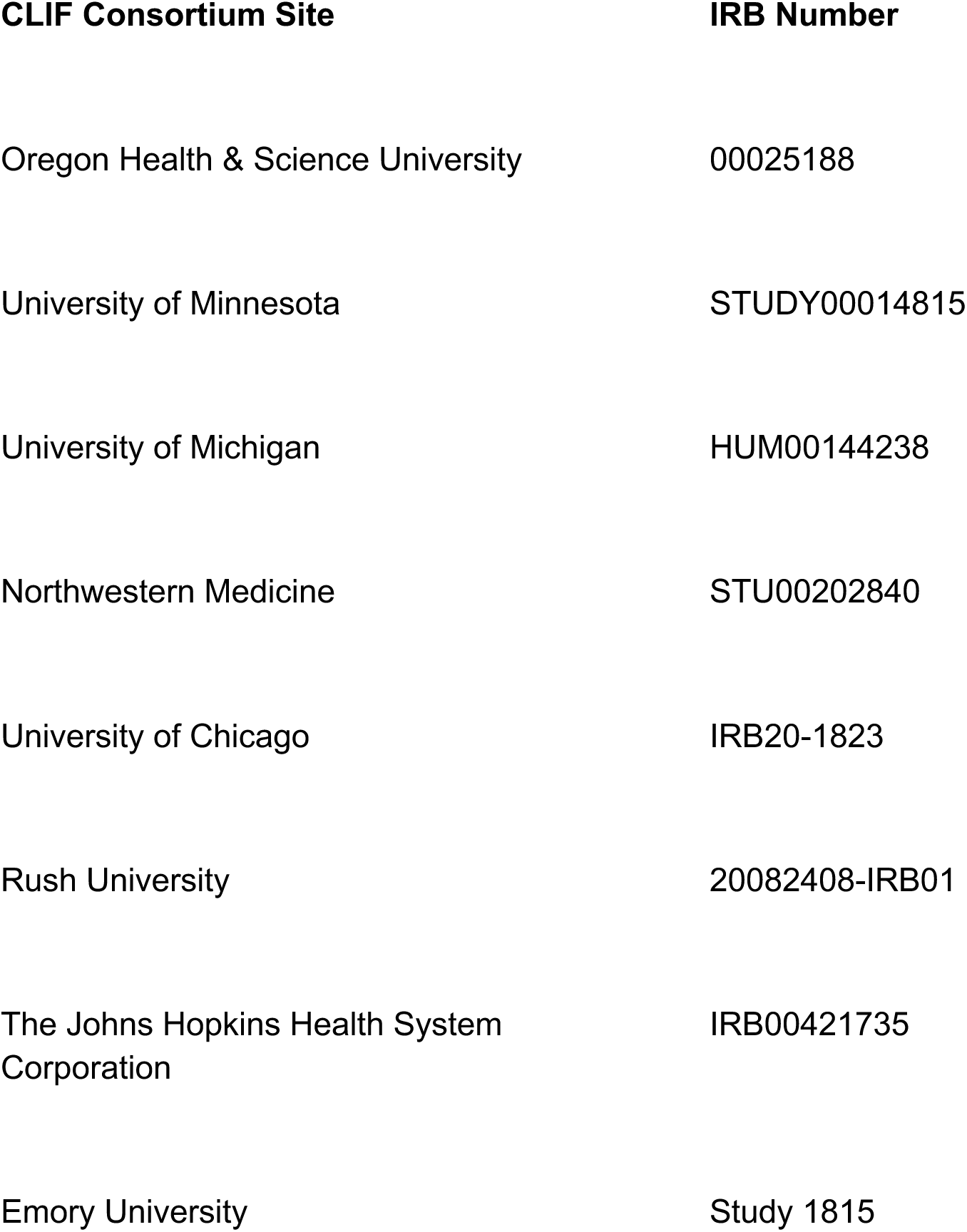
IRB Details for CLIF Consortium Sites. This supplement provides IRB approval numbers for observational studies of critically ill patients and EDW building/quality-checking activities conducted at CLIF consortium sites.

**Table E2:**
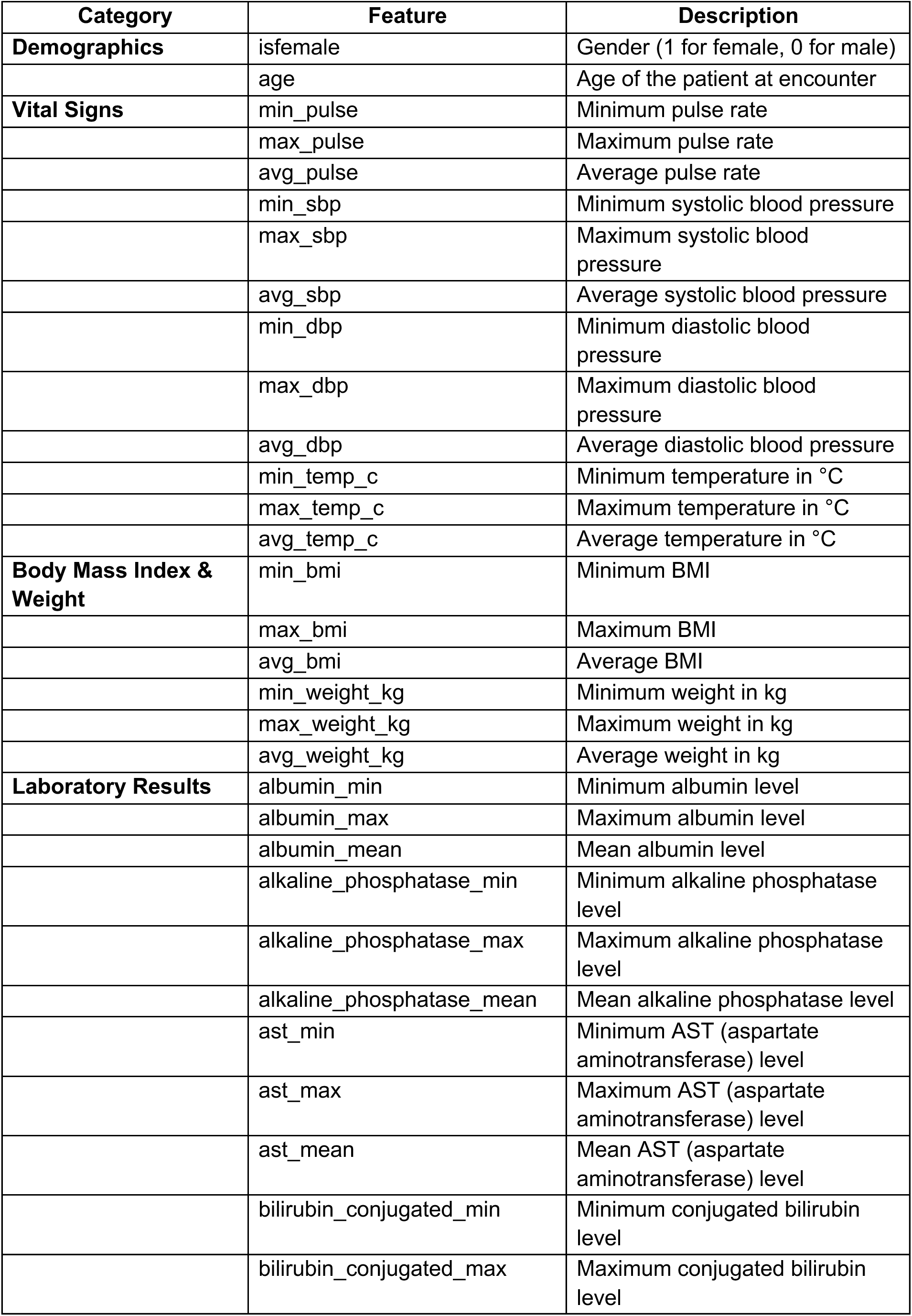
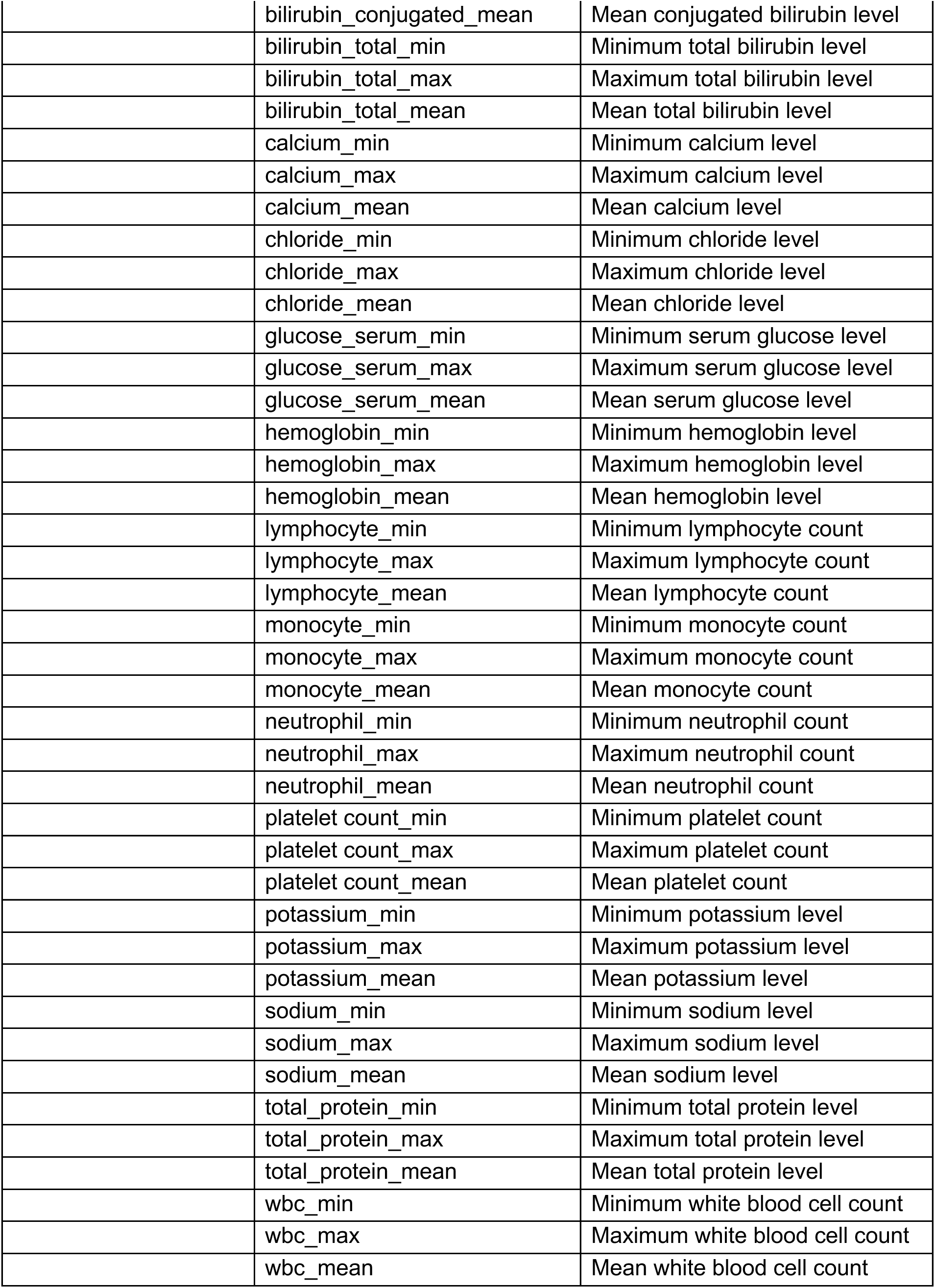
Features of the Machine Learning Model Predicting ICU Mortality During the First 24 Hours.

**Table E3:**
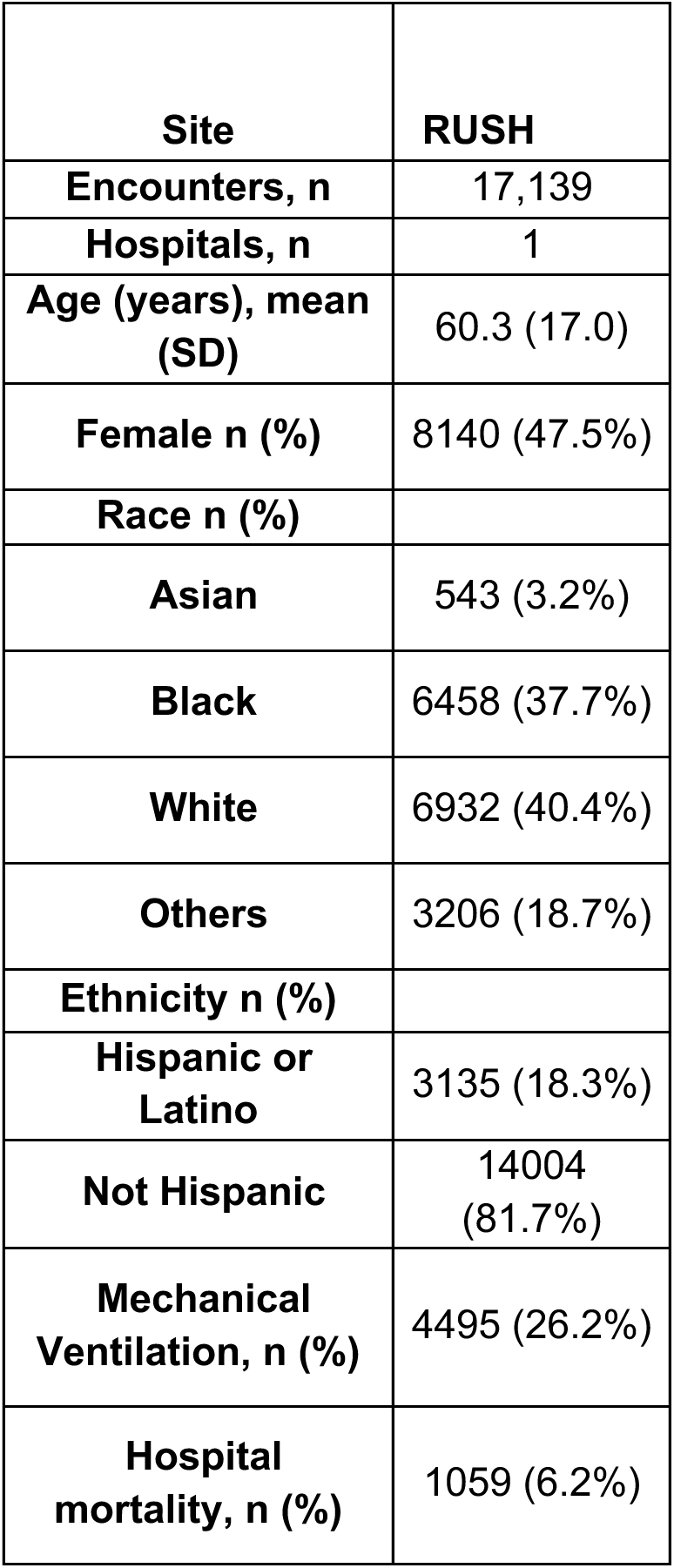
Derivation Cohort Characteristics for Inpatient Mortality Model at Rush University.

**Table E4:**
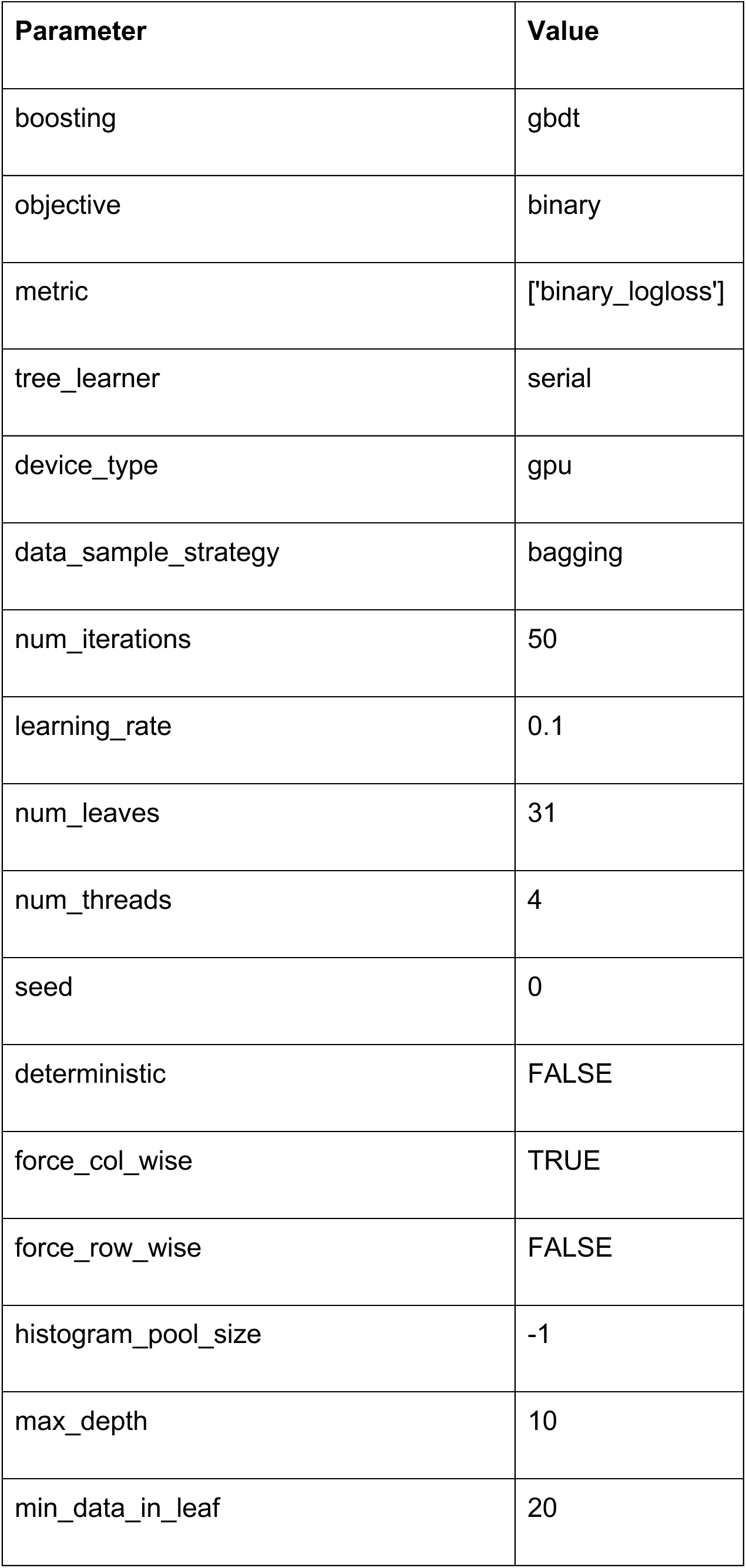
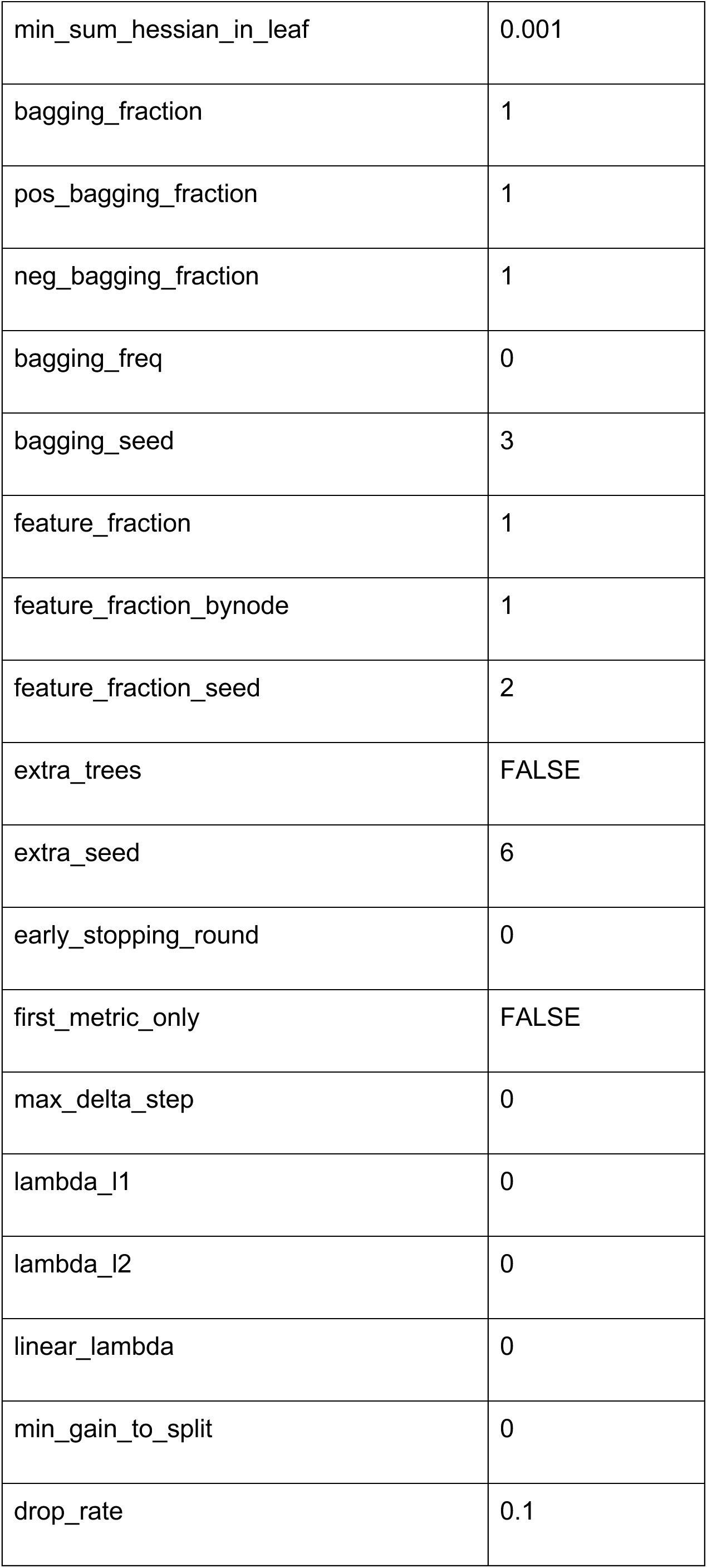
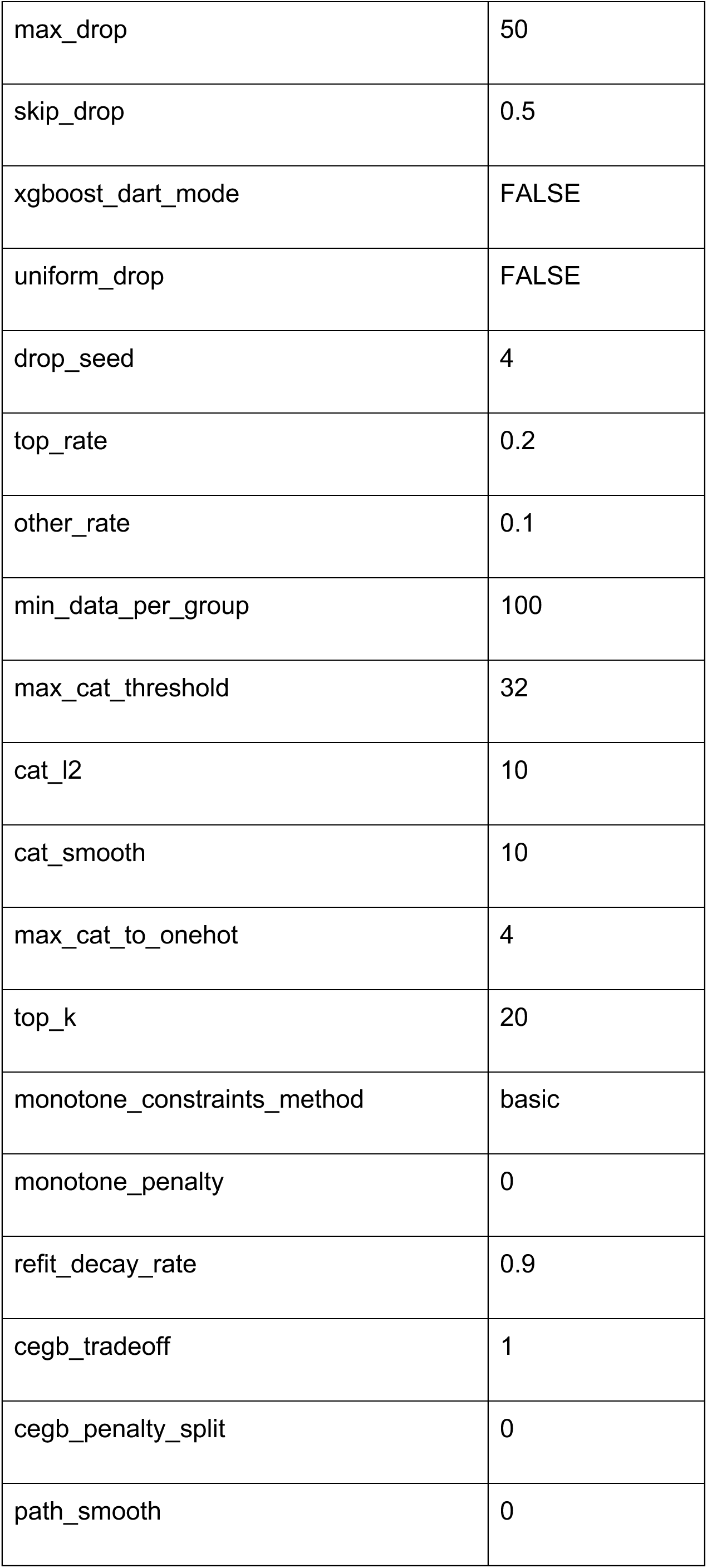
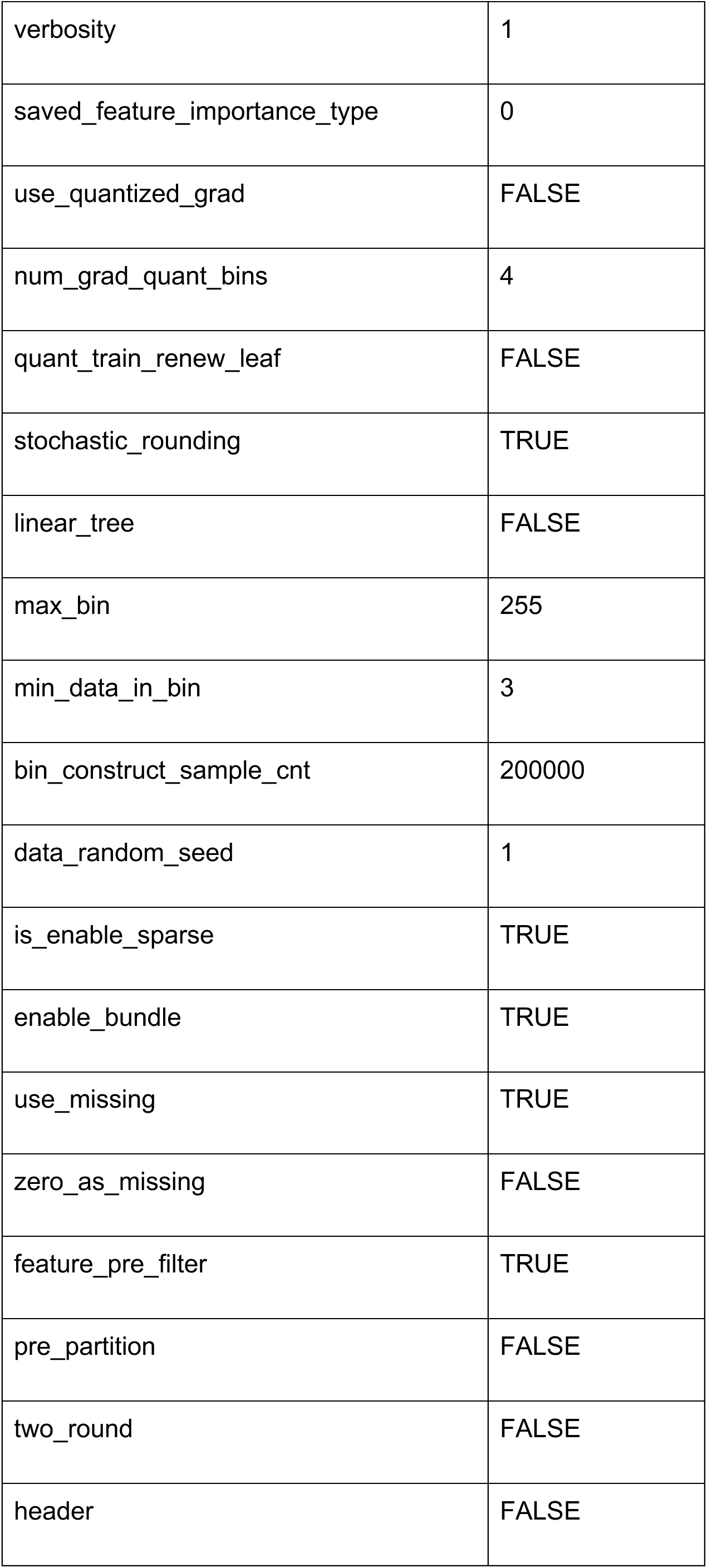
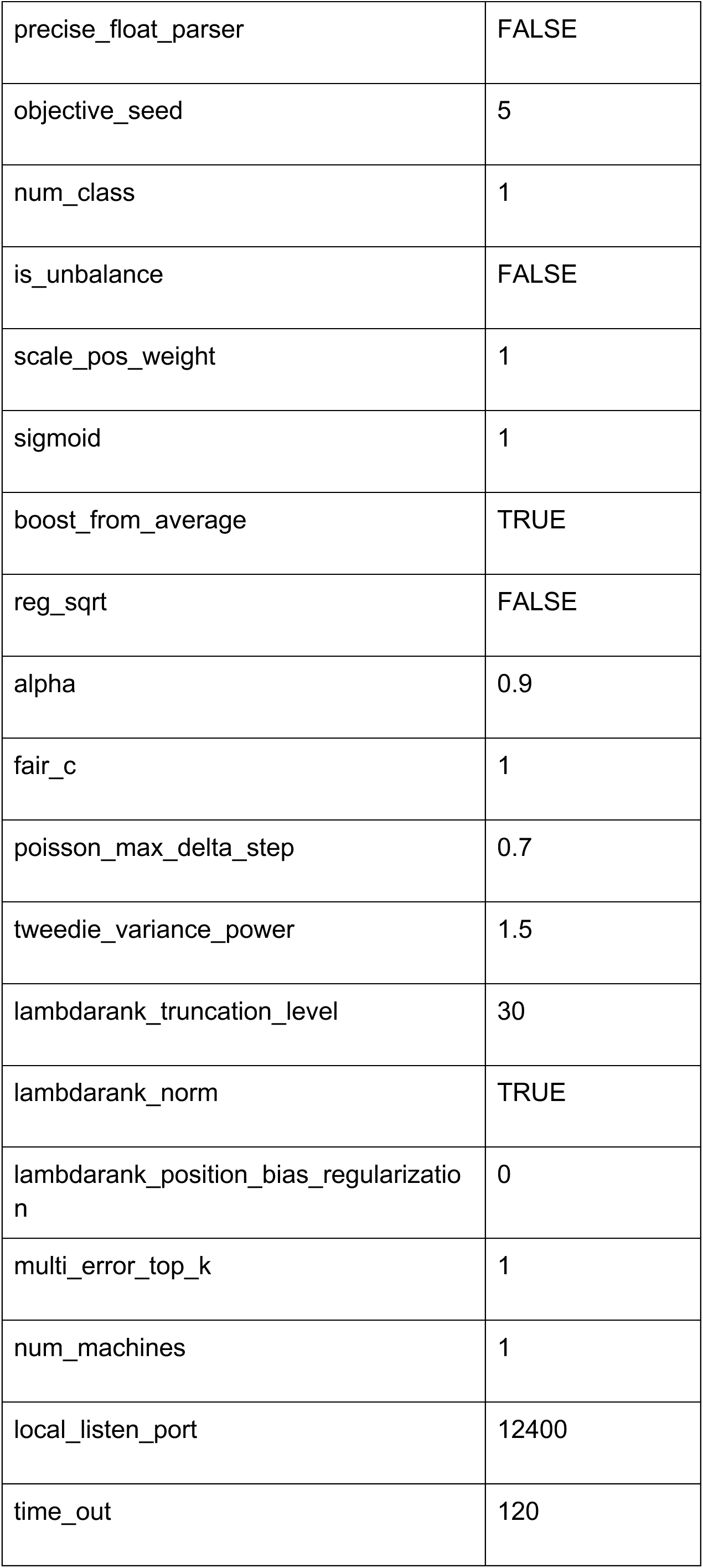
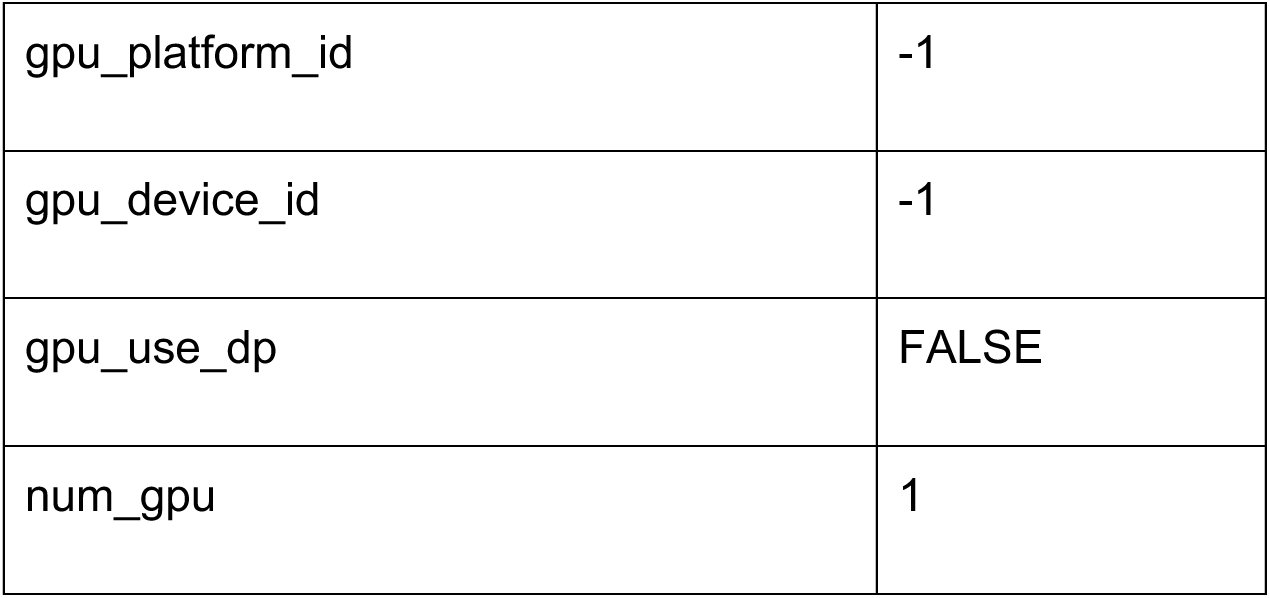
Hyperparameters for Inpatient Mortality LightGBM Model Derived at Rush.

**Table E5.**
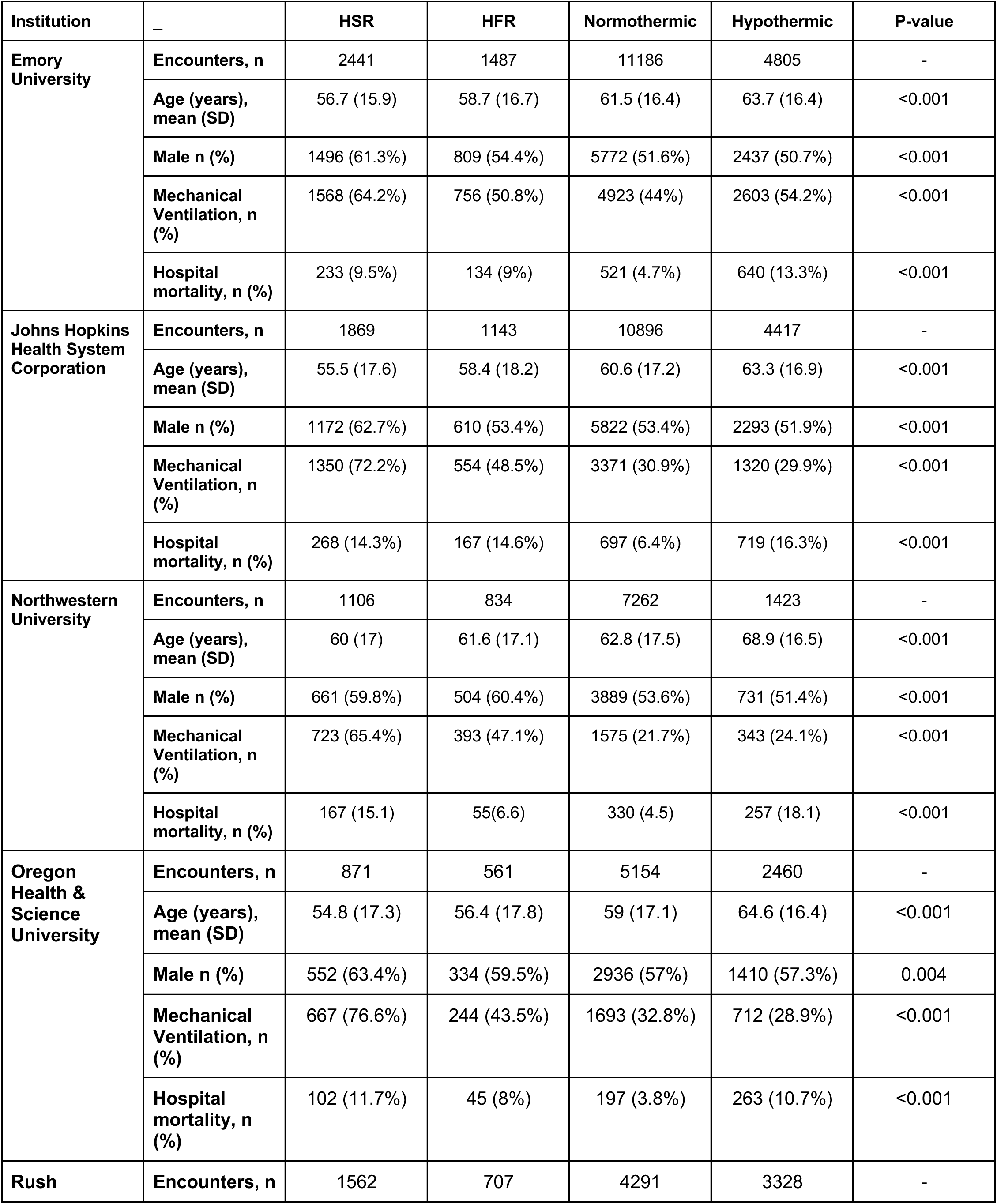
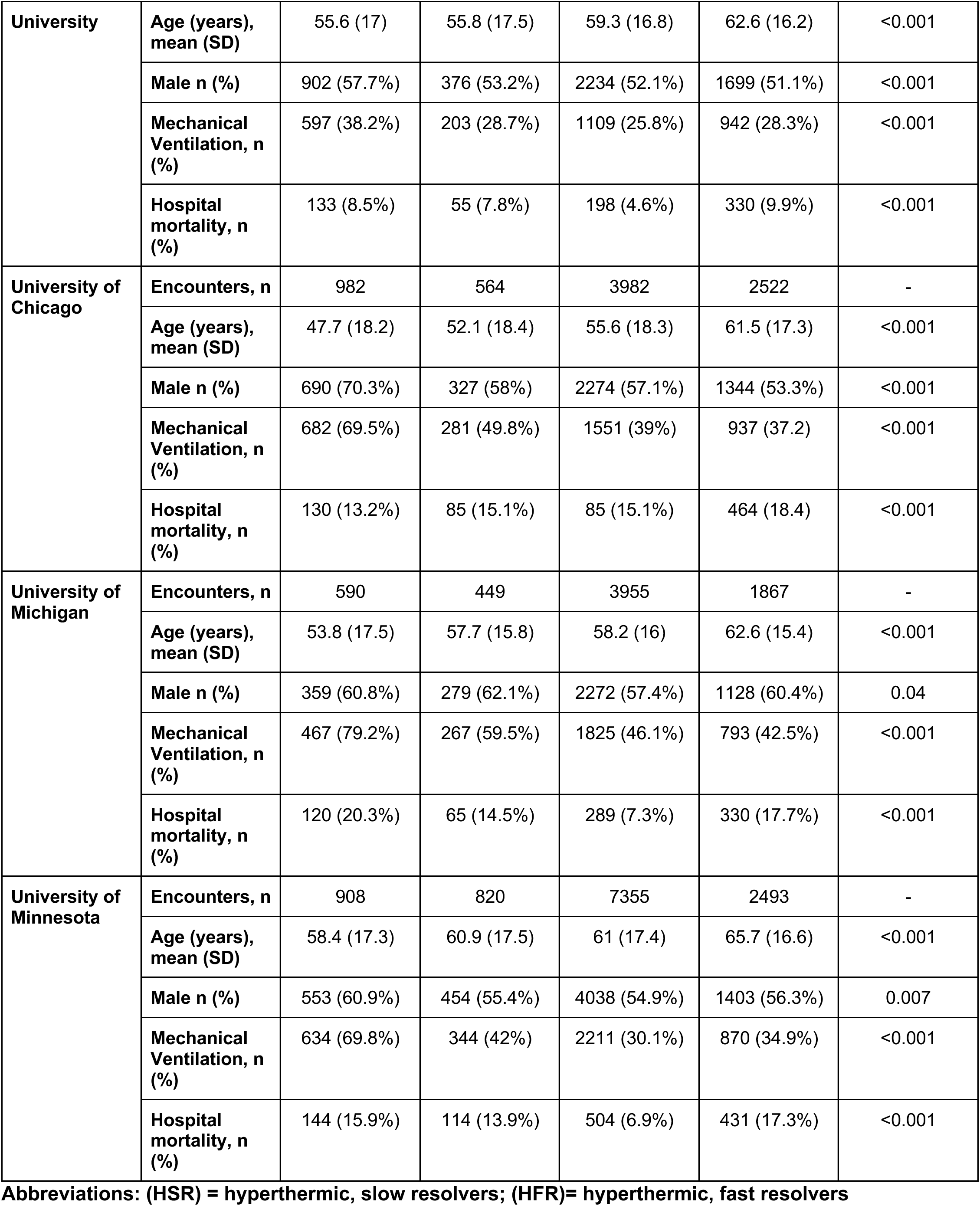
Temperature Trajectory Data by CLIF Site.

## Supplement Figure Legend

**Figure E1:**
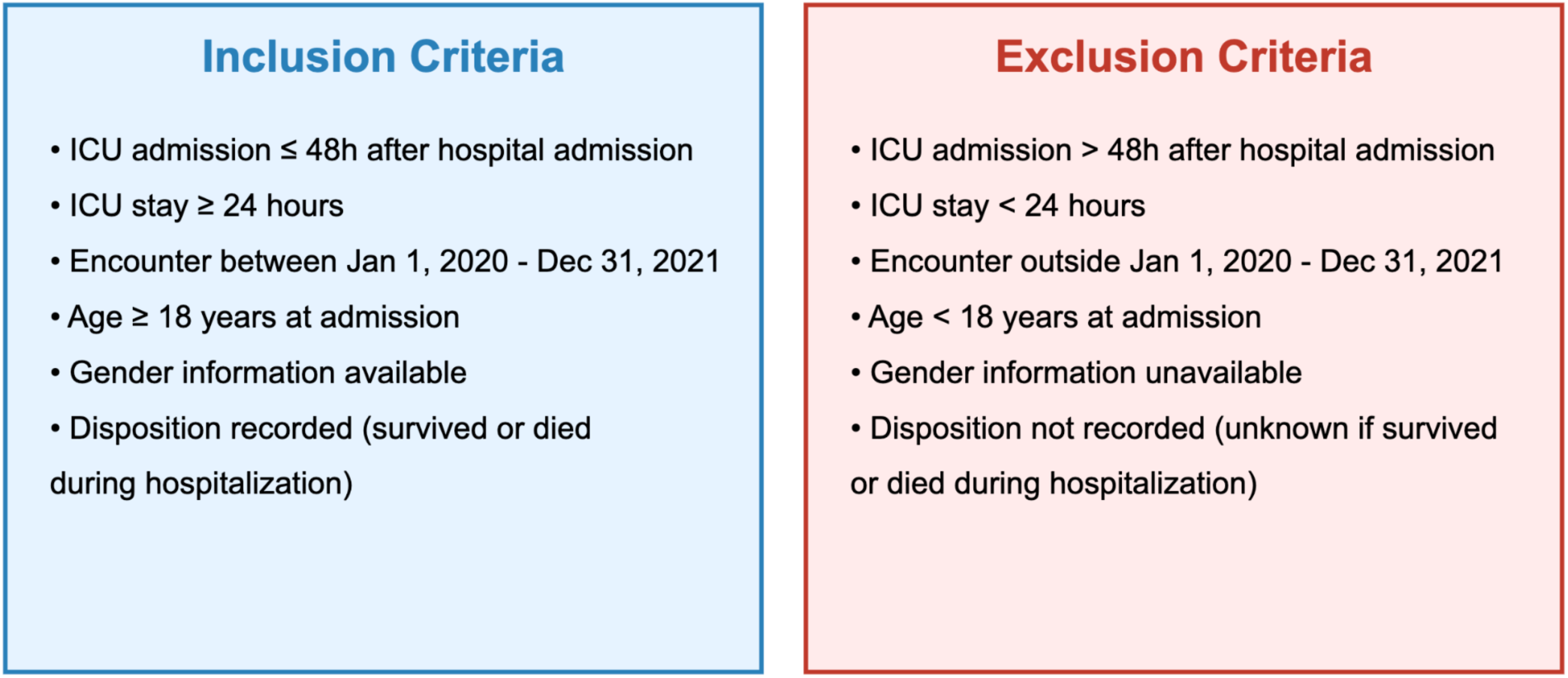
Inclusion and Exclusion Criteria for ICU CLIF Case Studies. This figure illustrates the detailed criteria used for including and excluding cases in the ICU CLIF case studies.

**Figure E2:**
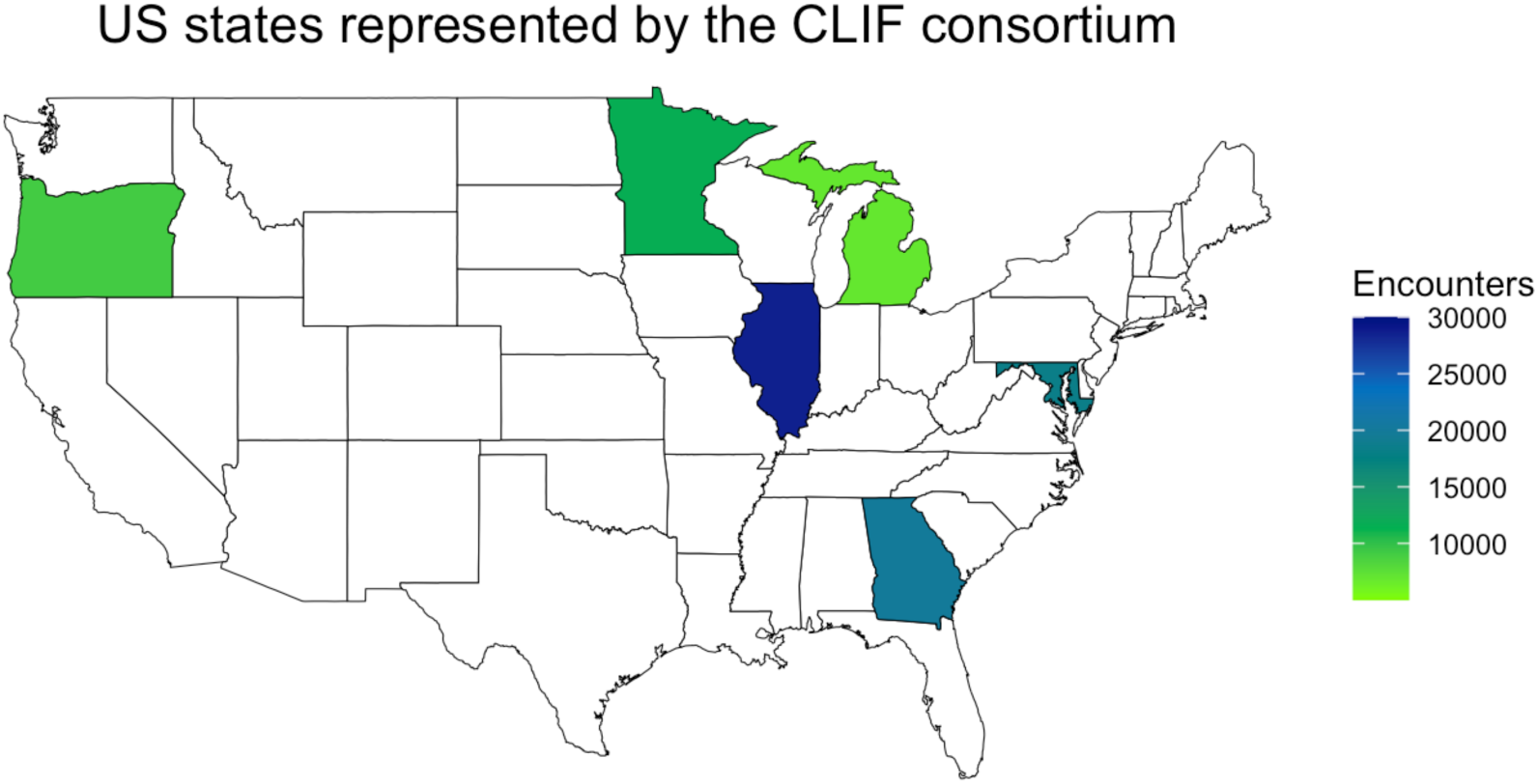
US States Represented by CLIF. This map highlights the US states represented in the CLIF study, with participating institutions listed alongside their respective states. The institutions include: Oregon Health & Science University (Oregon) University of Minnesota (Minnesota) University of Michigan (Michigan) Northwestern Medicine (Illinois) UChicago Medicine (Illinois) Rush University (Illinois) Johns Hopkins Medicine (Maryland) Emory University (Georgia)

**Figure E3:**
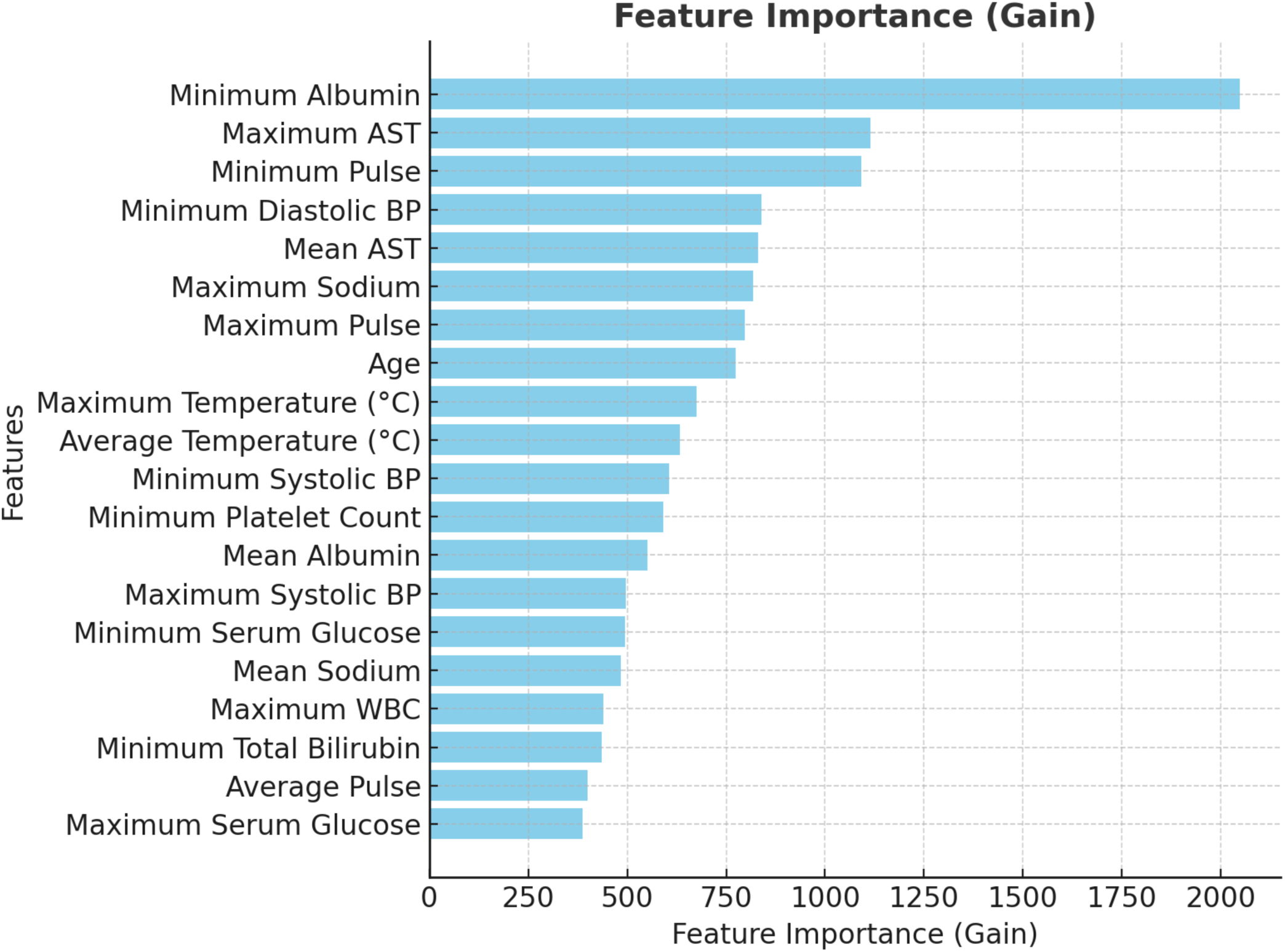
Variable Importance Plot for Inpatient Mortality Prediction Model. Feature Importance (Gain): This plot ranks the features (variables) in the model based on their significance in predicting inpatient mortality. The importance of each feature is measured using a metric called "Gain," which reflects how much each feature improves the model’s accuracy in predicting the likelihood of a patient’s survival during hospitalization. Gain: The higher the Gain for a feature, the more it contributes to the model’s ability to predict the target outcome, which in this case is inpatient mortality. Gain measures the impact of each feature on the decision-making process of the model, with higher Gain values indicating a greater influence on predicting whether a patient will survive or not. For example, if "Minimum Albumin" has the highest Gain value in the plot, it means that this feature is the most important for the model’s predictions of inpatient mortality. This implies that lower levels of albuminare strongly associated with a higher risk of mortality in hospitalized patients, more so than other measured variables. Legend: WBC: White Blood Cell count AST: Aspartate Aminotransferase, an enzyme found in the liver and other tissues BP: Blood Pressure

## REFERENCES

1. Johnson AEW, Pollard TJ, Shen L, et al. MIMIC-III, a freely accessible critical care database. Sci Data [Internet] 2016;3:160035. Available from: 10.1038/sdata.2016.35

2. Pollard TJ, Johnson AEW, Raffa JD, Celi LA, Mark RG, Badawi O. The eICU Collaborative Research Database, a freely available multi-center database for critical care research. Sci Data [Internet] 2018;5:180178. Available from: 10.1038/sdata.2018.178

3. Thoral PJ, Peppink JM, Driessen RH, et al. Sharing ICU Patient Data Responsibly Under the Society of Critical Care Medicine/European Society of Intensive Care Medicine Joint Data Science Collaboration: The Amsterdam University Medical Centers Database (AmsterdamUMCdb) Example. Crit Care Med [Internet] 2021;49(6):e563–77. Available from: 10.1097/CCM.0000000000004916

4. Rojas JC, Carey KA, Edelson DP, Venable LR, Howell MD, Churpek MM. Predicting Intensive Care Unit Readmission with Machine Learning Using Electronic Health Record Data. Ann Am Thorac Soc [Internet] 2018;15(7):846–53. Available from: 10.1513/AnnalsATS.201710-787OC

5. Nemati S, Holder A, Razmi F, Stanley MD, Clifford GD, Buchman TG. An Interpretable Machine Learning Model for Accurate Prediction of Sepsis in the ICU. Crit Care Med [Internet] 2018;46(4):547–53. Available from: 10.1097/CCM.0000000000002936

6. Hersh WR, Weiner MG, Embi PJ, et al. Caveats for the use of operational electronic health record data in comparative effectiveness research. Med Care [Internet] 2013;51(8 Suppl 3):S30–7. Available from: 10.1097/MLR.0b013e31829b1dbd

7. Denney MJ, Long DM, Armistead MG, Anderson JL, Conway BN. Validating the extract, transform, load process used to populate a large clinical research database. Int J Med Inform [Internet] 2016;94:271–4. Available from: 10.1016/j.ijmedinf.2016.07.009

8. Sun H, Depraetere K, De Roo J, et al. Semantic processing of EHR data for clinical research. J Biomed Inform [Internet] 2015;58:247–59. Available from: 10.1016/j.jbi.2015.10.009

9. Pedrera-Jiménez M, García-Barrio N, Cruz-Rojo J, et al. Obtaining EHR-derived datasets for COVID-19 research within a short time: a flexible methodology based on Detailed Clinical Models. J Biomed Inform [Internet] 2021;115:103697. Available from: 10.1016/j.jbi.2021.103697

10. Ong T, Pradhananga R, Holve E, Kahn MG. A Framework for Classification of Electronic Health Data Extraction-Transformation-Loading Challenges in Data Network Participation. EGEMS (Wash DC) [Internet] 2017;5(1):10. Available from: 10.5334/egems.222

11. Callahan A, Shah NH, Chen JH. Research and Reporting Considerations for Observational Studies Using Electronic Health Record Data. Ann Intern Med [Internet] 2020;172(11 Suppl):S79–84. Available from: 10.7326/M19-0873

12. Paris N, Lamer A, Parrot A. Transformation and Evaluation of the MIMIC Database in the OMOP Common Data Model: Development and Usability Study. JMIR Med Inform [Internet] 2021;9(12):e30970. Available from: 10.2196/30970

13. Huser V, Kahn MG, Brown JS, Gouripeddi R. Methods for examining data quality in healthcare integrated data repositories. Pac Symp Biocomput [Internet] 2018;23:628–33. Available from: https://www.ncbi.nlm.nih.gov/pubmed/29218922

14. Leese P, Anand A, Girvin A, et al. Clinical encounter heterogeneity and methods for resolving in networked EHR data: a study from N3C and RECOVER programs. J Am Med Inform Assoc [Internet] 2023;30(6):1125–36. Available from: 10.1093/jamia/ocad057

15. Data Management and Sharing Policy [Internet]. [cited 2024 Aug 28];Available from: https://sharing.nih.gov/data-management-and-sharing-policy

16. NIH Common Data Elements (CDE) repository [Internet]. [cited 2024 Aug 26];Available from: https://cde.nlm.nih.gov/home

17. Adams MCB, Hurley RW, Siddons A, Topaloglu U, Wandner LD. NIH HEAL clinical data elements (CDE) implementation: NIH HEAL Initiative IMPOWR network IDEA-CC. Pain Med [Internet] 2023;24(7):743–9. Available from: https://academic.oup.com/painmedicine/article-abstract/24/7/743/7043983

18. Bhavani SV, Carey KA, Gilbert ER, Afshar M, Verhoef PA, Churpek MM. Identifying Novel Sepsis Subphenotypes Using Temperature Trajectories. Am J Respir Crit Care Med [Internet] 2019;200(3):327–35. Available from: 10.1164/rccm.201806-1197OC

19. Lyons PG, Hofford MR, Yu SC, et al. Factors Associated With Variability in the Performance of a Proprietary Sepsis Prediction Model Across 9 Networked Hospitals in the US. JAMA Intern Med [Internet] 2023;183(6):611–2. Available from: 10.1001/jamainternmed.2022.7182

20. Lambden S, Laterre PF, Levy MM, Francois B. The SOFA score-development, utility and challenges of accurate assessment in clinical trials. Crit Care [Internet] 2019;23(1):374. Available from: 10.1186/s13054-019-2663-7

21. Miller WD, Han X, Peek ME, Charan Ashana D, Parker WF. Accuracy of the Sequential Organ Failure Assessment Score for In-Hospital Mortality by Race and Relevance to Crisis Standards of Care. JAMA Netw Open [Internet] 2021;4(6):e2113891. Available from: 10.1001/jamanetworkopen.2021.13891

22. Ashana DC, Anesi GL, Liu VX, et al. Equitably Allocating Resources during Crises: Racial Differences in Mortality Prediction Models. Am J Respir Crit Care Med [Internet] 2021;204(2):178–86. Available from: 10.1164/rccm.202012-4383OC

23. Ashana DC, Welsh W, Preiss D, et al. Racial Differences in Shared Decision-Making About Critical Illness. JAMA Intern Med [Internet] 2024;Available from: 10.1001/jamainternmed.2023.8433

24. Ke G, Meng Q, Finley T, et al. Lightgbm: A highly efficient gradient boosting decision tree. Adv Neural Inf Process Syst [Internet] 2017;30. Available from: https://proceedings.neurips.cc/paper/2017/hash/6449f44a102fde848669bdd9eb6b76fa-Abstract.html

25. Collins GS, Dhiman P, Andaur Navarro CL, et al. Protocol for development of a reporting guideline (TRIPOD-AI) and risk of bias tool (PROBAST-AI) for diagnostic and prognostic prediction model studies based on artificial intelligence. BMJ Open [Internet] 2021;11(7):e048008. Available from: 10.1136/bmjopen-2020-048008

26. Vickers AJ, Elkin EB. Decision curve analysis: a novel method for evaluating prediction models. Med Decis Making [Internet] 2006;26(6):565–74. Available from: 10.1177/0272989X06295361

27. Lyons PG, Meyer NJ, Maslove DM. The Road to Precision in Critical Care. Crit Care Med [Internet] 2024;Available from: 10.1097/CCM.0000000000006213

28. Bos LDJ, Sjoding M, Sinha P, et al. Longitudinal respiratory subphenotypes in patients with COVID-19-related acute respiratory distress syndrome: results from three observational cohorts. Lancet Respir Med [Internet] 2021;9(12):1377–86. Available from: 10.1016/S2213-2600(21)00365-9

29. Bhavani SV, Huang ES, Verhoef PA, Churpek MM. Novel Temperature Trajectory Subphenotypes in COVID-19. Chest [Internet] 2020;158(6):2436–9. Available from: 10.1016/j.chest.2020.07.027

30. Bhavani SV, Verhoef PA, Maier CL, et al. Coronavirus Disease 2019 Temperature Trajectories Correlate With Hyperinflammatory and Hypercoagulable Subphenotypes. Crit Care Med [Internet] 2022;50(2):212–23. Available from: 10.1097/CCM.0000000000005397

31. Benzoni NS, Carey KA, Bewley AF, et al. Temperature Trajectory Subphenotypes in Oncology Patients with Neutropenia and Suspected Infection. Am J Respir Crit Care Med [Internet] 2023;207(10):1300–9. Available from: 10.1164/rccm.202205-0920OC

32. Kaissis GA, Makowski MR, Rückert D, Braren RF. Secure, privacy-preserving and federated machine learning in medical imaging. Nat Mach Intell [Internet] 2020;2(6):305–11. Available from: https://www.nature.com/articles/s42256-020-0186-1

33. Bongers KS, Chanderraj R, Woods RJ, et al. The Gut Microbiome Modulates Body Temperature Both in Sepsis and Health. Am J Respir Crit Care Med [Internet] 2023;207(8):1030–41. Available from: 10.1164/rccm.202201-0161OC

34. Brat GA, Weber GM, Gehlenborg N, et al. International electronic health record-derived COVID-19 clinical course profiles: the 4CE consortium. NPJ Digit Med [Internet] 2020;3:109. Available from: 10.1038/s41746-020-00308-0

35. Weiskopf NG, Dorr DA, Jackson C, Lehmann HP, Thompson CA. Healthcare utilization is a collider: an introduction to collider bias in EHR data reuse. J Am Med Inform Assoc [Internet] 2023;30(5):971–7. Available from: 10.1093/jamia/ocad013

36. Johnson AEW, Bulgarelli L, Shen L, et al. MIMIC-IV, a freely accessible electronic health record dataset. Sci Data [Internet] 2023;10(1):1. Available from: 10.1038/s41597-022-01899-x

37. critical [Internet]. [cited 2024 Aug 28];Available from: https://critical-consortium.github.io/

