## Supplement for "A Common Longitudinal Intensive Care Unit data Format (CLIF) to enable multi-institutional federated critical illness research"

Running Title: CLIF: Standardizing ICU Data for Federated Research

**Table E1: IRB Details for CLIF Consortium Sites**

This supplement provides IRB approval numbers for observational studies of critically ill patients and EDW building/quality-checking activities conducted at CLIF consortium sites.

| CLIF Consortium Site | IRB Number |
|---|---|
| Oregon Health & Science University | 00025188 |
| University of Minnesota | STUDY00014815 |
| University of Michigan | HUM00144238 |
| Northwestern Medicine | STU00202840 |
| University of Chicago | IRB20-1823 |
| Rush University | 20082408-IRB01 |
| The Johns Hopkins Health System Corporation | IRB00421735 |
| Emory University | Study 1815 |

**Table E2: Features of the Machine Learning Model Predicting ICU Mortality During the First 24 Hours**

| Category | Feature | Description |
|---|---|---|
| **Demographics** | isfemale | Gender (1 for female, 0 for male) |
| | age | Age of the patient at encounter |
| **Vital Signs** | min_pulse | Minimum pulse rate |
| | max_pulse | Maximum pulse rate |
| | avg_pulse | Average pulse rate |
| | min_sbp | Minimum systolic blood pressure |
| | max_sbp | Maximum systolic blood pressure |
| | avg_sbp | Average systolic blood pressure |
| | min_dbp | Minimum diastolic blood pressure |
| | max_dbp | Maximum diastolic blood pressure |
| | avg_dbp | Average diastolic blood pressure |
| | min_temp_c | Minimum temperature in °C |
| | max_temp_c | Maximum temperature in °C |
| | avg_temp_c | Average temperature in °C |
| **Body Mass Index & Weight** | min_bmi | Minimum BMI |
| | max_bmi | Maximum BMI |
| | avg_bmi | Average BMI |
| | min_weight_kg | Minimum weight in kg |
| | max_weight_kg | Maximum weight in kg |
| | avg_weight_kg | Average weight in kg |
| **Laboratory Results** | albumin_min | Minimum albumin level |
| | albumin_max | Maximum albumin level |
| | albumin_mean | Mean albumin level |
| | alkaline_phosphatase_min | Minimum alkaline phosphatase level |
| | alkaline_phosphatase_max | Maximum alkaline phosphatase level |
| | alkaline_phosphatase_mean | Mean alkaline phosphatase level |
| | ast_min | Minimum AST (aspartate aminotransferase) level |
| | ast_max | Maximum AST (aspartate aminotransferase) level |
| | ast_mean | Mean AST (aspartate aminotransferase) level |
| | bilirubin_conjugated_min | Minimum conjugated bilirubin level |
| | bilirubin_conjugated_max | Maximum conjugated bilirubin level |

| | | |
|---|---|---|
| | bilirubin_conjugated_mean | Mean conjugated bilirubin level |
| | bilirubin_total_min | Minimum total bilirubin level |
| | bilirubin_total_max | Maximum total bilirubin level |
| | bilirubin_total_mean | Mean total bilirubin level |
| | calcium_min | Minimum calcium level |
| | calcium_max | Maximum calcium level |
| | calcium_mean | Mean calcium level |
| | chloride_min | Minimum chloride level |
| | chloride_max | Maximum chloride level |
| | chloride_mean | Mean chloride level |
| | glucose_serum_min | Minimum serum glucose level |
| | glucose_serum_max | Maximum serum glucose level |
| | glucose_serum_mean | Mean serum glucose level |
| | hemoglobin_min | Minimum hemoglobin level |
| | hemoglobin_max | Maximum hemoglobin level |
| | hemoglobin_mean | Mean hemoglobin level |
| | lymphocyte_min | Minimum lymphocyte count |
| | lymphocyte_max | Maximum lymphocyte count |
| | lymphocyte_mean | Mean lymphocyte count |
| | monocyte_min | Minimum monocyte count |
| | monocyte_max | Maximum monocyte count |
| | monocyte_mean | Mean monocyte count |
| | neutrophil_min | Minimum neutrophil count |
| | neutrophil_max | Maximum neutrophil count |
| | neutrophil_mean | Mean neutrophil count |
| | platelet count_min | Minimum platelet count |
| | platelet count_max | Maximum platelet count |
| | platelet count_mean | Mean platelet count |
| | potassium_min | Minimum potassium level |
| | potassium_max | Maximum potassium level |
| | potassium_mean | Mean potassium level |
| | sodium_min | Minimum sodium level |
| | sodium_max | Maximum sodium level |
| | sodium_mean | Mean sodium level |
| | total_protein_min | Minimum total protein level |
| | total_protein_max | Maximum total protein level |
| | total_protein_mean | Mean total protein level |
| | wbc_min | Minimum white blood cell count |
| | wbc_max | Maximum white blood cell count |
| | wbc_mean | Mean white blood cell count |

**Table E3: Derivation Cohort Characteristics for Inpatient Mortality Model at Rush University**

| Site | RUSH |
|---|---|
| Encounters, n | 17,139 |
| Hospitals, n | 1 |
| Age (years), mean (SD) | 60.3 (17.0) |
| Female n (%) | 8140 (47.5%) |
| Race n (%) | |
| Asian | 543 (3.2%) |
| Black | 6458 (37.7%) |
| White | 6932 (40.4%) |
| Others | 3206 (18.7%) |
| Ethnicity n (%) | |
| Hispanic or Latino | 3135 (18.3%) |
| Not Hispanic | 14004 (81.7%) |
| Mechanical Ventilation, n (%) | 4495 (26.2%) |
| Hospital mortality, n (%) | 1059 (6.2%) |

**Table E4: Hyperparameters for Inpatient Mortality LightGBM Model Derived at Rush**

| Parameter | Value |
|---|---|
| boosting | gbdt |
| objective | binary |
| metric | ['binary_logloss'] |
| tree_learner | serial |
| device_type | gpu |
| data_sample_strategy | bagging |
| num_iterations | 50 |
| learning_rate | 0.1 |
| num_leaves | 31 |
| num_threads | 4 |
| seed | 0 |
| deterministic | FALSE |
| force_col_wise | TRUE |
| force_row_wise | FALSE |
| histogram_pool_size | -1 |
| max_depth | 10 |
| min_data_in_leaf | 20 |

| | |
|---|---|
| min_sum_hessian_in_leaf | 0.001 |
| bagging_fraction | 1 |
| pos_bagging_fraction | 1 |
| neg_bagging_fraction | 1 |
| bagging_freq | 0 |
| bagging_seed | 3 |
| feature_fraction | 1 |
| feature_fraction_bynode | 1 |
| feature_fraction_seed | 2 |
| extra_trees | FALSE |
| extra_seed | 6 |
| early_stopping_round | 0 |
| first_metric_only | FALSE |
| max_delta_step | 0 |
| lambda_l1 | 0 |
| lambda_l2 | 0 |
| linear_lambda | 0 |
| min_gain_to_split | 0 |
| drop_rate | 0.1 |

| | |
|---|---|
| max_drop | 50 |
| skip_drop | 0.5 |
| xgboost_dart_mode | FALSE |
| uniform_drop | FALSE |
| drop_seed | 4 |
| top_rate | 0.2 |
| other_rate | 0.1 |
| min_data_per_group | 100 |
| max_cat_threshold | 32 |
| cat_l2 | 10 |
| cat_smooth | 10 |
| max_cat_to_onehot | 4 |
| top_k | 20 |
| monotone_constraints_method | basic |
| monotone_penalty | 0 |
| refit_decay_rate | 0.9 |
| cegb_tradeoff | 1 |
| cegb_penalty_split | 0 |
| path_smooth | 0 |

| | |
|---|---|
| verbosity | 1 |
| saved_feature_importance_type | 0 |
| use_quantized_grad | FALSE |
| num_grad_quant_bins | 4 |
| quant_train_renew_leaf | FALSE |
| stochastic_rounding | TRUE |
| linear_tree | FALSE |
| max_bin | 255 |
| min_data_in_bin | 3 |
| bin_construct_sample_cnt | 200000 |
| data_random_seed | 1 |
| is_enable_sparse | TRUE |
| enable_bundle | TRUE |
| use_missing | TRUE |
| zero_as_missing | FALSE |
| feature_pre_filter | TRUE |
| pre_partition | FALSE |
| two_round | FALSE |
| header | FALSE |

| | |
|---|---|
| precise_float_parser | FALSE |
| objective_seed | 5 |
| num_class | 1 |
| is_unbalance | FALSE |
| scale_pos_weight | 1 |
| sigmoid | 1 |
| boost_from_average | TRUE |
| reg_sqrt | FALSE |
| alpha | 0.9 |
| fair_c | 1 |
| poisson_max_delta_step | 0.7 |
| tweedie_variance_power | 1.5 |
| lambdarank_truncation_level | 30 |
| lambdarank_norm | TRUE |
| lambdarank_position_bias_regularization | 0 |
| multi_error_top_k | 1 |
| num_machines | 1 |
| local_listen_port | 12400 |
| time_out | 120 |

| | |
|---|---|
| gpu_platform_id | -1 |
| gpu_device_id | -1 |
| gpu_use_dp | FALSE |
| num_gpu | 1 |

### Table E5. Temperature Trajectory Data by CLIF Site

| Institution | _ | HSR | HFR | Normothermic | Hypothermic | P-value |
|---|---|---|---|---|---|---|
| **Emory University** | **Encounters, n** | 2441 | 1487 | 11186 | 4805 | - |
| | **Age (years), mean (SD)** | 56.7 (15.9) | 58.7 (16.7) | 61.5 (16.4) | 63.7 (16.4) | <0.001 |
| | **Male n (%)** | 1496 (61.3%) | 809 (54.4%) | 5772 (51.6%) | 2437 (50.7%) | <0.001 |
| | **Mechanical Ventilation, n (%)** | 1568 (64.2%) | 756 (50.8%) | 4923 (44%) | 2603 (54.2%) | <0.001 |
| | **Hospital mortality, n (%)** | 233 (9.5%) | 134 (9%) | 521 (4.7%) | 640 (13.3%) | <0.001 |
| **Johns Hopkins Health System Corporation** | **Encounters, n** | 1869 | 1143 | 10896 | 4417 | - |
| | **Age (years), mean (SD)** | 55.5 (17.6) | 58.4 (18.2) | 60.6 (17.2) | 63.3 (16.9) | <0.001 |
| | **Male n (%)** | 1172 (62.7%) | 610 (53.4%) | 5822 (53.4%) | 2293 (51.9%) | <0.001 |
| | **Mechanical Ventilation, n (%)** | 1350 (72.2%) | 554 (48.5%) | 3371 (30.9%) | 1320 (29.9%) | <0.001 |
| | **Hospital mortality, n (%)** | 268 (14.3%) | 167 (14.6%) | 697 (6.4%) | 719 (16.3%) | <0.001 |
| **Northwestern University** | **Encounters, n** | 1106 | 834 | 7262 | 1423 | - |
| | **Age (years), mean (SD)** | 60 (17) | 61.6 (17.1) | 62.8 (17.5) | 68.9 (16.5) | <0.001 |
| | **Male n (%)** | 661 (59.8%) | 504 (60.4%) | 3889 (53.6%) | 731 (51.4%) | <0.001 |
| | **Mechanical Ventilation, n (%)** | 723 (65.4%) | 393 (47.1%) | 1575 (21.7%) | 343 (24.1%) | <0.001 |
| | **Hospital mortality, n (%)** | 167 (15.1) | 55(6.6) | 330 (4.5) | 257 (18.1) | <0.001 |
| **Oregon Health & Science University** | **Encounters, n** | 871 | 561 | 5154 | 2460 | - |
| | **Age (years), mean (SD)** | 54.8 (17.3) | 56.4 (17.8) | 59 (17.1) | 64.6 (16.4) | <0.001 |
| | **Male n (%)** | 552 (63.4%) | 334 (59.5%) | 2936 (57%) | 1410 (57.3%) | 0.004 |
| | **Mechanical Ventilation, n (%)** | 667 (76.6%) | 244 (43.5%) | 1693 (32.8%) | 712 (28.9%) | <0.001 |
| | **Hospital mortality, n (%)** | 102 (11.7%) | 45 (8%) | 197 (3.8%) | 263 (10.7%) | <0.001 |
| **Rush** | **Encounters, n** | 1562 | 707 | 4291 | 3328 | - |

| | | | | | | |
|---|---|---|---|---|---|---|
| **University** | **Age (years), mean (SD)** | 55.6 (17) | 55.8 (17.5) | 59.3 (16.8) | 62.6 (16.2) | <0.001 |
| | **Male n (%)** | 902 (57.7%) | 376 (53.2%) | 2234 (52.1%) | 1699 (51.1%) | <0.001 |
| | **Mechanical Ventilation, n (%)** | 597 (38.2%) | 203 (28.7%) | 1109 (25.8%) | 942 (28.3%) | <0.001 |
| | **Hospital mortality, n (%)** | 133 (8.5%) | 55 (7.8%) | 198 (4.6%) | 330 (9.9%) | <0.001 |
| **University of Chicago** | **Encounters, n** | 982 | 564 | 3982 | 2522 | - |
| | **Age (years), mean (SD)** | 47.7 (18.2) | 52.1 (18.4) | 55.6 (18.3) | 61.5 (17.3) | <0.001 |
| | **Male n (%)** | 690 (70.3%) | 327 (58%) | 2274 (57.1%) | 1344 (53.3%) | <0.001 |
| | **Mechanical Ventilation, n (%)** | 682 (69.5%) | 281 (49.8%) | 1551 (39%) | 937 (37.2) | <0.001 |
| | **Hospital mortality, n (%)** | 130 (13.2%) | 85 (15.1%) | 85 (15.1%) | 464 (18.4) | <0.001 |
| **University of Michigan** | **Encounters, n** | 590 | 449 | 3955 | 1867 | - |
| | **Age (years), mean (SD)** | 53.8 (17.5) | 57.7 (15.8) | 58.2 (16) | 62.6 (15.4) | <0.001 |
| | **Male n (%)** | 359 (60.8%) | 279 (62.1%) | 2272 (57.4%) | 1128 (60.4%) | 0.04 |
| | **Mechanical Ventilation, n (%)** | 467 (79.2%) | 267 (59.5%) | 1825 (46.1%) | 793 (42.5%) | <0.001 |
| | **Hospital mortality, n (%)** | 120 (20.3%) | 65 (14.5%) | 289 (7.3%) | 330 (17.7%) | <0.001 |
| **University of Minnesota** | **Encounters, n** | 908 | 820 | 7355 | 2493 | - |
| | **Age (years), mean (SD)** | 58.4 (17.3) | 60.9 (17.5) | 61 (17.4) | 65.7 (16.6) | <0.001 |
| | **Male n (%)** | 553 (60.9%) | 454 (55.4%) | 4038 (54.9%) | 1403 (56.3%) | 0.007 |
| | **Mechanical Ventilation, n (%)** | 634 (69.8%) | 344 (42%) | 2211 (30.1%) | 870 (34.9%) | <0.001 |
| | **Hospital mortality, n (%)** | 144 (15.9%) | 114 (13.9%) | 504 (6.9%) | 431 (17.3%) | <0.001 |

**Abbreviations: (HSR) = hyperthermic, slow resolvers; (HFR)= hyperthermic, fast resolvers**

**Supplement Figure Legend:**

**Figure E1: Inclusion and Exclusion Criteria for ICU CLIF Case Studies**
This figure illustrates the detailed criteria used for including and excluding cases in the ICU CLIF case studies.

**Figure E2: US States Represented by CLIF**
This map highlights the US states represented in the CLIF study, with participating institutions listed alongside their respective states. The institutions include:
Oregon Health & Science University (Oregon)
University of Minnesota (Minnesota)
University of Michigan (Michigan)
Northwestern Medicine (Illinois)
UChicago Medicine (Illinois)
Rush University (Illinois)
Johns Hopkins Medicine (Maryland)
Emory University (Georgia)

**Figure E3: Variable Importance Plot for Inpatient Mortality Prediction Model**
Feature Importance (Gain): This plot ranks the features (variables) in the model based on their significance in predicting inpatient mortality. The importance of each feature is measured using a metric called "Gain," which reflects how much each feature improves the model's accuracy in predicting the likelihood of a patient's survival during hospitalization.
Gain: The higher the Gain for a feature, the more it contributes to the model's ability to predict the target outcome, which in this case is inpatient mortality. Gain measures the impact of each feature on the decision-making process of the model, with higher Gain values indicating a greater influence on predicting whether a patient will survive or not.
For example, if "Minimum Albumin" has the highest Gain value in the plot, it means that this feature is the most important for the model's predictions of inpatient mortality. This implies that lower levels of albuminare strongly associated with a higher risk of mortality in hospitalized patients, more so than other measured variables.

Legend:
WBC: White Blood Cell count
AST: Aspartate Aminotransferase, an enzyme found in the liver and other tissues
BP: Blood Pressure

**Figure E1: Inclusion and Exclusion Criteria for ICU CLIF Case Studies**

### Inclusion Criteria

- ICU admission ≤ 48h after hospital admission
- ICU stay ≥ 24 hours
- Encounter between Jan 1, 2020 - Dec 31, 2021
- Age ≥ 18 years at admission
- Gender information available
- Disposition recorded (survived or died during hospitalization)

### Exclusion Criteria

- ICU admission > 48h after hospital admission
- ICU stay < 24 hours
- Encounter outside Jan 1, 2020 - Dec 31, 2021
- Age < 18 years at admission
- Gender information unavailable
- Disposition not recorded (unknown if survived or died during hospitalization)

**Figure E2. US States represented by CLIF**

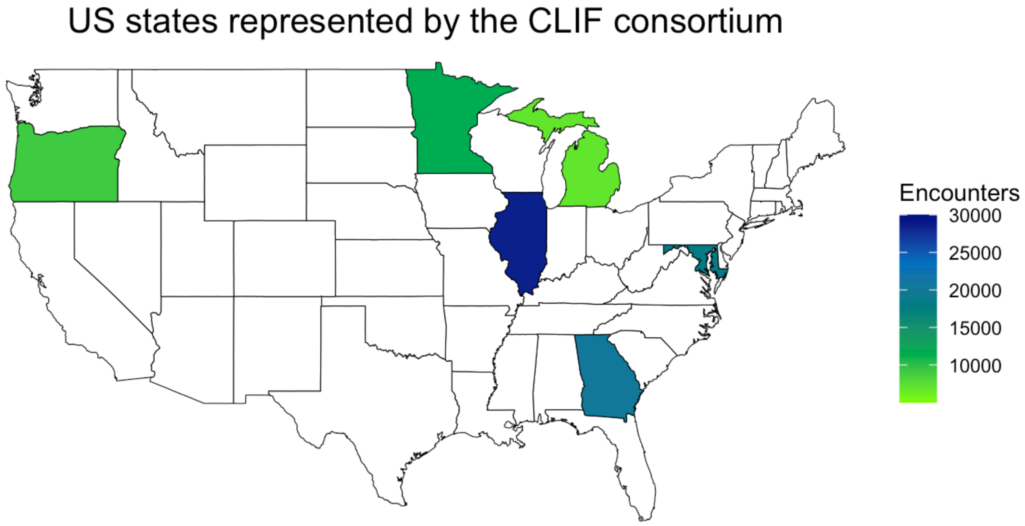

**Figure E3: Variable Importance Plot for Inpatient Mortality Prediction Model**

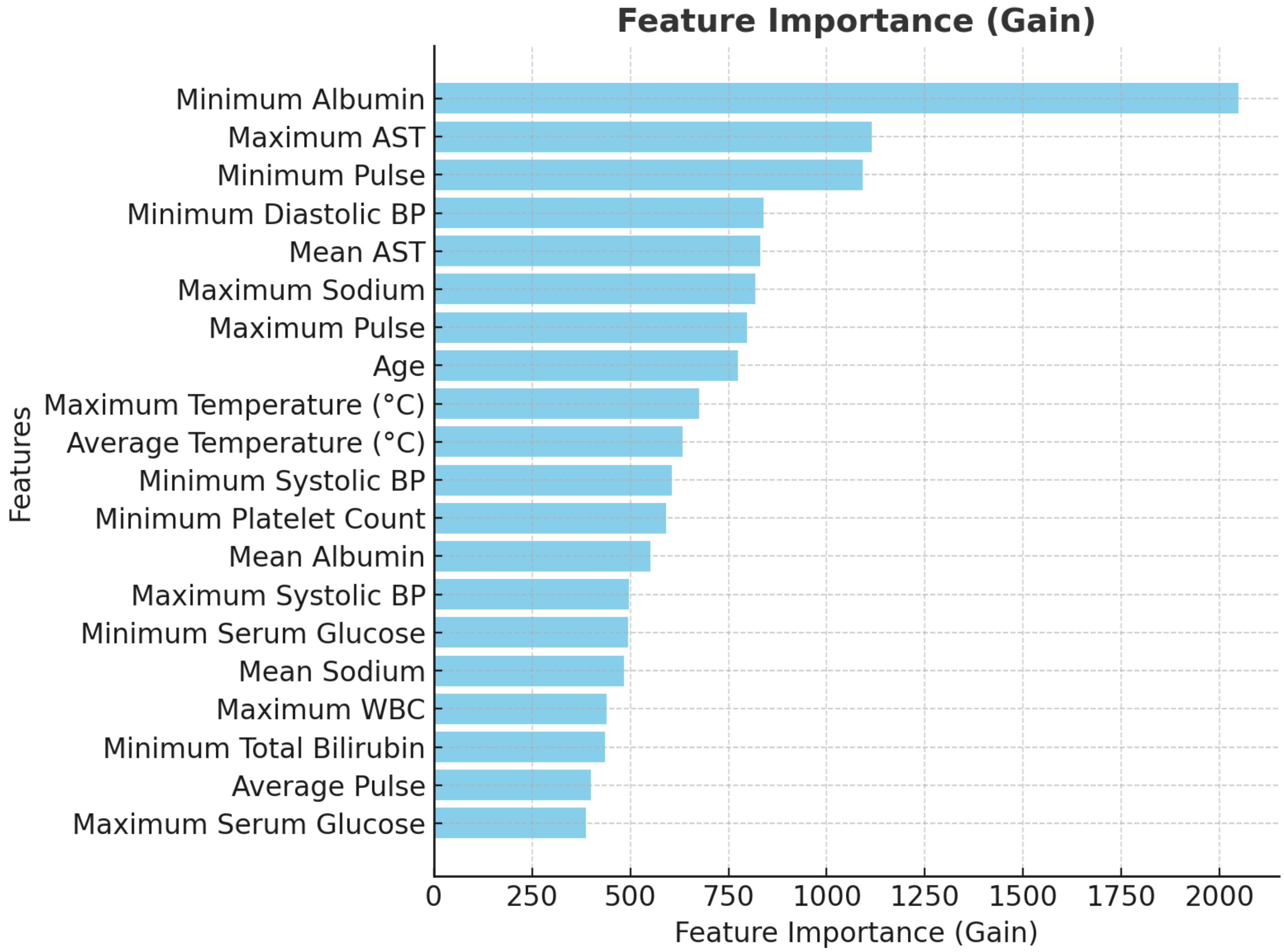
